# M-PreSS: A Model Pre-training Approach for Study Screening in Systematic Reviews

**DOI:** 10.1101/2025.04.08.25325463

**Authors:** Zhaozhen Xu, Philippa Davies, Louise AC Millard, Lam Teng, Georgios Markozannes, Pau Erola, Eduardo AP Seleiro, Julian PT Higgins, Richard M Martin, Maria Sobczyk-Barad, Konstantinos K Tsilidis, Doris SM Chan, Tom R Gaunt, Yi Liu

## Abstract

**Background:** Conducting a systematic review is labour-intensive and time-consuming, especially during the study screening process. Previous research has introduced traditional machine learning models (e.g. Support Vector Machines) to automate the study screening process, but it is difficult to generalise across topics. Therefore, recent research has explored the use of existing large language models (LLMs), such as ChatGPT/GPT-4, for study screening. However, the lack of transparency in training data and consistency in output results make applying such commercial LLMs challenging in the context of systematic reviews where transparency in methods is particularly important.

**Results:** We introduce an approach to fine-tune an open-source biomedical language model (BlueBERT) using a Siamese neural network [1] so that it screens the scientific literature databases on multiple research topics. We evaluate different training approaches in seven COVID-19 systematic reviews. The results indicate good generalisation among topics with an average recall/sensitivity of 0.86 (minimum: 0.67, maximum: 1.00) and an average false positive rate of 6.48% (minimum: 1.38%, maximum: 11.41%). Furthermore, adding study selection criteria to the topic definition can improve the model performance (Area Under the Precision-Recall Curve [PRAUC]) by 2.74%, and adding more related review topics during training can increase the performance by 15.82%.

**Conclusions:** Our results indicate that fine-tuning BlueBERT with study screening datasets can outperform ChatGPT/GPT-4 in two out of three COVID-19 review topics reported in the literature, whilst maintaining the ability for researchers to continue updating or extending the search for related evidence and significantly reducing the computational resource requirements.

## 1. Introduction

Systematic literature reviews are widely used to support evidence-based research and inform policy and practice in many domains, including biomedical and clinical sciences. Compared to a narrative literature review, which gives a summary or overview of a topic, a systematic review also aims to provide a transparent, thorough, and unbiased evaluation of all the existing evidence for a specific research question, including the assessment of the risk of bias, and often the quantitative summation of study results using a meta-analysis. Guidelines [2] are provided for conducting systematic reviews, which aim to make the processes transparent and repeatable. The review team might want to update the review with new evidence or answer different questions under a general research theme, where reproducibility and efficiency can be improved by the use of computational tools [3].

Automation in systematic reviews has been widely studied due to the potential for computerising the repetitive systematic processes involved. The process to automate usually breaks down into various tasks following different stages of conducting systematic reviews, such as search of study, study screening, data extraction, risk of bias assessment to evaluate the validity of the study results, evidence synthesis, evidence quality assessment, and reporting [4]. In this study, we focus on one of these key processes: study screening.

Study screening (also known as study selection) aims to identify the primary studies for a review topic (i.e. a specific research question). To include all related studies, the traditional approach is to conduct an exhaustive search in relevant bibliographic databases using pre-defined searching strategies, followed by manual screening of the retrieved study records according to the eligibility criteria. In practice, a search from literature databases can return thousands of candidate studies for each systematic review topic. Then, a minimum of two reviewers typically independently screen the study records and agree on a final decision on whether a study should be included or excluded in the systematic review [5]. The screening typically comprises two phases. In the initial phase, the titles and abstracts are screened to remove obviously irrelevant references, which significantly reduces the number of references for the next phase. Then, the full-text papers are screened to make the decisions on inclusion in the review. The decisions are recorded during the process.

A significant challenge in study screening is the substantial time and human resources needed. A previous study [6] surveyed 33 experienced systematic review reviewers and found that it can take a full-time systematic reviewer an average of 33 days for the entire screening process. Hence, automating the study screening process with machine learning (ML) has the potential to increase the efficiency of conducting systematic reviews.

Recent research [7, 8, 9] apply ML and natural language processing (NLP) approaches to train models that can automate the manual screening process with the existing study screening records from published systematic reviews. However, these trained models cannot be generalised among different review questions nor extended to new review topics. Some studies [10, 11] leverage ChatGPT/GPT-4 for study screening without any training on the screening records and demonstrate that the model can exhibit good generalisation on new review topics. Nevertheless, concerns regarding the model’s lack of transparency, such as the training data and methodology, and consistency in output results remain, which is critical in the context of systematic review.

In this study, we introduce M-PreSS, a **m**odel **pre**-training approach for **s**tudy **s**creening, for different systematic review topics. This approach aims to train a general study model that can be used among multiple topics and potentially extended to new/unseen topics. A general model is not expected to excel in every topic but demonstrates robust performance across a broad range of topics. For instance, a model is general when it performs well on multiple cancer-related topics, even if it does not achieve good performance across all disease categories.

Our model is based on BlueBERT [12], a biomedical version of the original BERT network. BERT [13] is a well-established large language model (LLM) pre-trained with Wikipedia and BookCorpus, and demonstrates strong performance across various language understanding tasks. Based on the original BERT, BlueBERT is further trained with PubMed abstracts and clinical notes, which results in capturing biomedicine language patterns and performance in multiple biomedical NLP tasks, such as sentence similarity and named entity recognition. By fine-tuning BlueBERT with existing study screening datasets, the model may then be more effectively directed at identifying relevant studies on the given systematic review topic.

Moreover, our study leverages a Siamese network [1] to fine-tune BlueBERT so that one general model can be used across topics. The Siamese network is an architecture that consists of two identical subnetworks for comparing the relationship and similarity between two inputs [1]. In our study, the Siamese network allows the model to take both review topic and reference as inputs, and make different inclusion/exclusion decisions for a reference depending on the topic provided. Siamese networks have been shown to be robust to imbalanced data for classification task [14]. Another study [15] has also demonstrated that Siamese structures can achieve higher recall for positive examples (i.e. included studies) compared to cross-encoders, which concatenate inputs for training. Therefore, it is applicable for study screening where there are few positive examples and identifying all relevant studies is important (i.e. high recall).50 Combining the power of LLM and Siamese networks, M-PreSS can be generalised to multiple topics (e.g. research questions) for a research focus (e.g. effcacy studies of COVID-19), which can be useful for research teams consistently looking for different types of evidence for a particular focus. More details of the M-PreSS architecture are explained in Section 3.

Section 4 presents the methodology for training and evaluating the M-PreSS model. When training our model, we leverage screening datasets from multiple sources. The idea is for the model to capture the general skill for study screening from diverse topics so that it can improve the overall performance among these topics, and can be potentially generalised to unseen topics. We train our model with various data sizes and topic homogeneity and evaluate its performance on open datasets to understand the impact of different training data on model performance. Moreover, to further explore the model’s generalisability, we evaluate it on unseen topics. The results are demonstrated in Section 5.

## 2. Related Work

Existing tools [16] and models [7, 8] apply traditional ML and NLP approaches to identify primary studies and have shown promise in using ML models to semi-automate study screening in the fields of software engineering, medicine, and biology [7, 8]. However, a specific ML model needs to be trained for each review topic to obtain optimal prediction results, and cannot then be directly applied to new topics without new labelled screening records from the reviewers and further training. Another challenge of applying ML to study screening is class imbalance. After screening, only a small percentage of candidate studies are included in the review, which limits the amount of ‘positive’ examples in the training data. This can cause a bias towards the majority class during training, which limits the performance of the learnt model on identifying relevant studies. Previous research [8] observed that up-sampling of included studies resulted in poor predictions and increasing computation time in study screening, while down-sampling of excluded studies was a better strategy for imbalanced data but resulted in reducing training data.

Recent developments in ML, particularly in LLMs, such as BERT [13] and GPTs [17, 18], have resulted in models that understand key aspects of human language and learn general knowledge by pre-training on large amounts of text, such as books, Wikipedia, and academic literature. Such models therefore have the potential to be used in various tasks related to the processing of natural language (e.g. information retrieval [19], text classification [20]) and for public health research problems [21, 22]. Furthermore, LLMs exhibit a good generalisation to unseen data without any training (i.e. zero-shot learning) and are increasingly used for automating systematic reviews [23].

One previous study [9] fine-tuned BERT to classify included and excluded studies using their titles and abstracts. Similar to traditional ML approaches, each systematic review topic requires its own model to be trained. Training a topic-specific screening model in this way relies on a sufficient number of human-labelled records, which would not typically be the case in real-world usage.

Some studies [10, 24, 25] have shown the potential to use an existing language model to perform zero-shot classification on different topics. However, they use pre-trained models without tailoring them for study screening. Thus, the recall and false positive rate of those models can potentially be improved by fine-tuning the model with existing study selection datasets.

Another study [11] demonstrates that using ChatGPT and GPT-4 can provide promising results as a complementary tool to human reviewers in screening clinical reviews. However, GPT-4 misclassified 22.8% of the relevant studies (average recall: 0.77) as irrelevant. In addition, responses from ChatGPT can be inconsistent across sessions, limiting the reproducibility of this approach. The results of previous studies highlight the need to investigate whether fine-tuning an open-source LLM with systematic review datasets can further improve the performance of the LLM on study screening. Our work builds on Guo et al.’s by fine-tuning a different LLM using the same data and comparing the performance with their results.

## 3. Method: General Model for Study Screening

We call M-PreSS a general model as it aims to use one model for multiple topics instead of customising topic-specific models. Figure 1 illustrates the architecture of it. M-Press produces a priority score for each candidate study based on the systematic review topic. During the initial study screening process, reviewers can utilise the trained model to rank studies and generate machine predictions on inclusion/exclusion, which support their decision-making.

**Figure 1:**
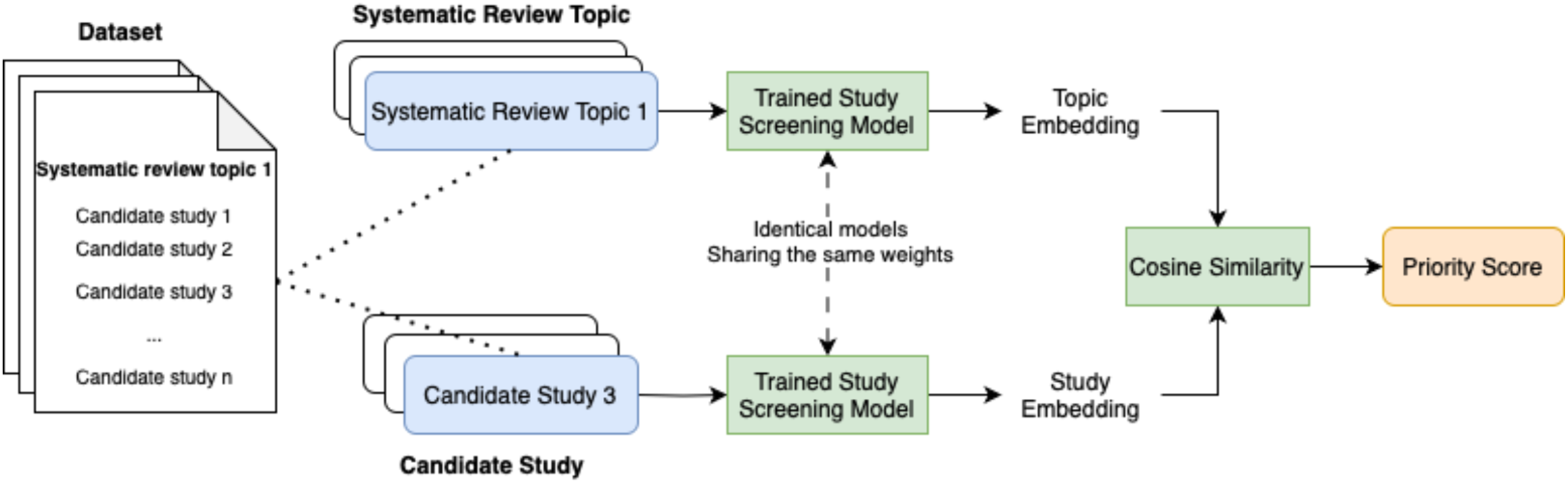
Architecture of M-PreSS. Blue: Input (*Systematic Review Topic, Candidate Study*) pair. Green: M-PreSS. Orange: Output priority score. Note that the trained study screening models are identical, which means they share the same setup and weights.

**Figure 2:**
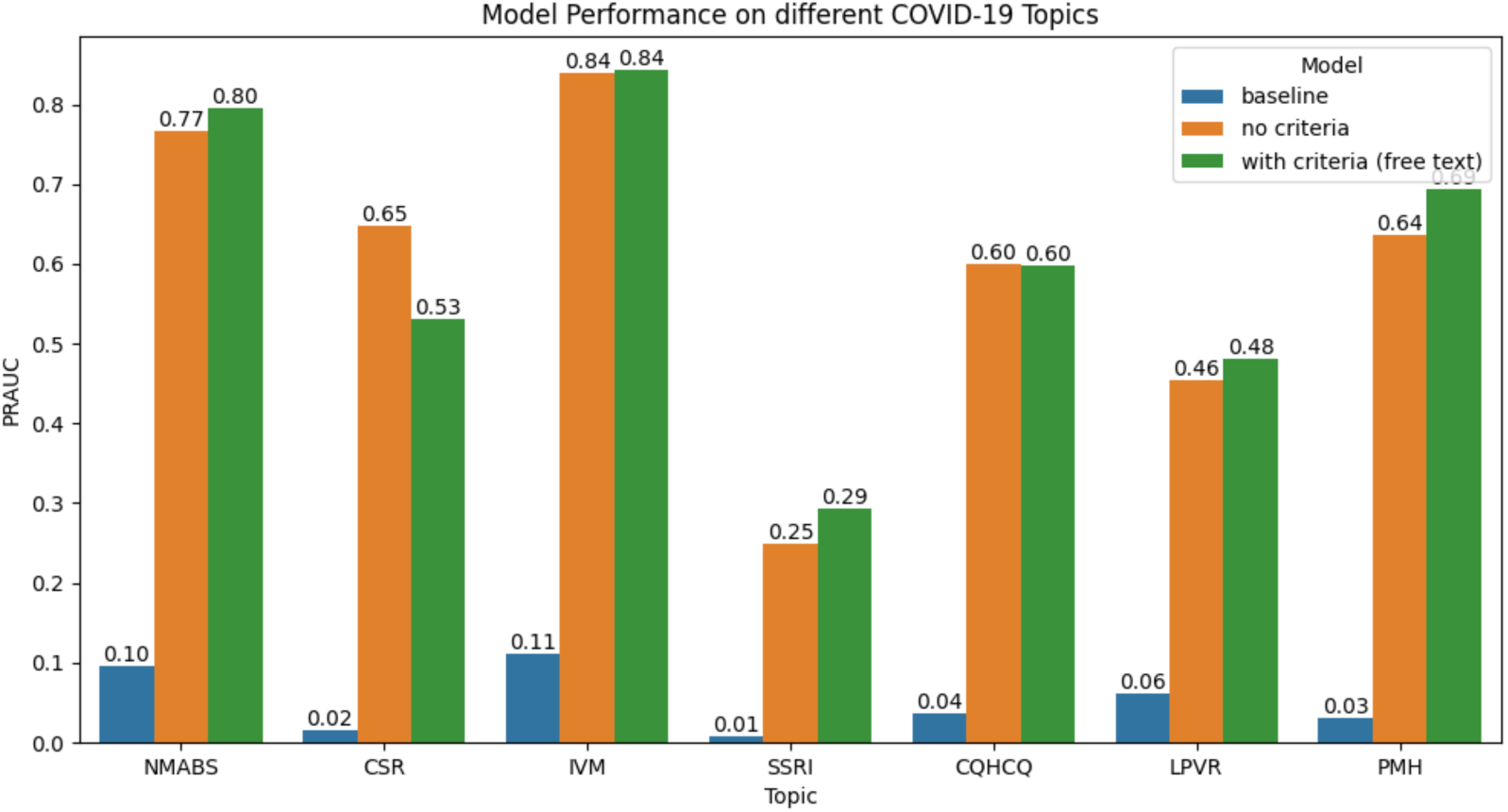
Comparing the models trained with and without selection criteria on the COVID-19 dataset. The criteria were manually extracted from the published systematic reviews. The models were trained on the training set and evaluated on the test set. The baseline for PRAUC were the inclusion percentage.

M-PreSS takes (*Systematic Review Topic, Candidate Study*) pairs as inputs, where the number of candidate studies per topic differs across the dataset. The training size is determined by the total number of candidate studies across all topics.

The systematic review topic is the published systematic review title. The candidate study includes the title and abstract. During training, some keywords, such as ‘Query’, ‘Title’, and ‘Abstract’, were added to the data to assist the language model in capturing the information in the topic and study. The input is constructed as below.

Query: {Systematic review title}.

Title: {Study title}. Abstract: {Study Abstract}

Then, the trained study model is able to embed the inputs into dense vector representations (called embeddings). These embeddings can capture the semantic meaning and key features of the topic and study.

The output of M-PreSS is a priority score, which can be used to rank the candidate studies for later review. The priority score is the cosine similarity between the systematic review topic and the candidate study embeddings. During the screening, the reviewers can pay less attention to studies with extremely high or low priority scores, thus increasing their efficiency.

Examples of input and output data from the model can be found in supplementary material Table A.6.

To train the study screening model, we employ a Siamese BERT structure using the Sentence-BERT [26] framework. Sentence-BERT leverages the Siamese architecture to capture the text similarity between two input sentences. The embeddings of sentences with similar meanings are close to each other in the embedding space.

In this study, we utilise the same Siamese structure to capture the relationship between the given systematic review topic and a candidate study. However, instead of the sentence-level inputs used in Sentence-BERT, we take the entire topic and candidate study as document-level inputs. We train the study screening model by fine-tuning BlueBERT using existing study screening records from previously completed systematic reviews. The aim is that a relevant study receives a higher cosine similarity to the topic than an irrelevant one during training.

Once trained, a topic and candidate study can be input and a priority score is assigned based on their relationship. Candidate studies relevant to the review topic are supposed to receive a higher priority score than irrelevant ones. The priority score for a given candidate study varies with the review topic, enabling one general model to determine whether a study should be included for various topics.

### 3.1. Binary Decision Threshold

When M-Press is used to assist human reviewers, it is also helpful for the model to propose a decision on whether a candidate study should be included in a specific review and a boundary for a reviewer to stop looking for relevant studies. Therefore, we introduce a decision threshold to give a binary output on inclusion/exclusion. If a (*Systematic Review Topic, Candidate Study*) pair has a priority score higher than the defined threshold, it is recommended for inclusion. The proposed decision can also be used to compare the performance of the trained model with that of human reviewers.

A specific threshold is calculated for each topic. As the inclusion/exclusion decision is local to a specific topic and the general priority score cannot be compared between topics, we calculated the topic-specific threshold based on the priority score and the ground truth for each topic to maximise the performance. Furthermore, due to the variability of text, such as language usage and semantic structures, an embedding distance that signifies relevance in one topic might not indicate the same level of relevance in another topic. Therefore, using a topic-specific threshold can accommodate this variability and increase classification accuracy.

The threshold is determined by ranking studies based on their priority score for the topic. Each score in this ordered list serves as a possible threshold. We can identify the optimal threshold from all the possible thresholds using the human labelling.

Multiple strategies can be used to identify the optimal thresholds, such as a threshold that results in the best accuracy metric on the training data. However, in the study screening task for a systematic review, the aim is to include all relevant studies. Therefore, we calculate the threshold that provides the lowest false positive rate (FPR) at 100% recall in the training set, encouraging the model to include all relevant studies without too many false positives for a human to examine.

More specifically, we rank the candidate studies for a topic by the priority score, and calculate the recall (Equation 1) and FPR (Equation 2) using each priority score as the threshold.

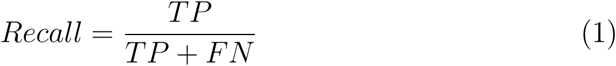

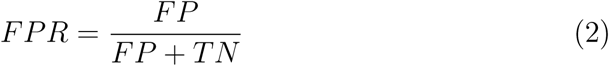

Where *TP*, *FN*, *FP*, and *TN* denote true positives, false negatives, false positives, and true negatives, respectively, from the confusion matrix [27]. The threshold is defined as the average of the priority score yielding the lowest FPR at 100% recall and the score of the next lower-ranked study. The process of identifying the best threshold is referred to as the ‘best FPR at 100% recall’ strategy and is demonstrated in Algorithm 1.

## 4. Experiments

To understand the performance and generalisability of our model, we trained the model on various data sources and different setups.

### 4.1. Datasets

We used existing systematic review screening records from different sources to train our model. Each review topic contained a list of study titles, abstracts, human decisions of inclusion or exclusion, and selection criteria.

*Guo et al.* [11] *(GUO).* Guo et al. provided screening records for ten clinical systematic reviews. All the topics were related to efficacy studies; out of them, seven studies are related to COVID-19. However, only three COVID-19 topics have inclusion/exclusion criteria and were evaluated in their study.

*Intervention Effectiveness.* We curated study screening records for six systematic reviews addressing questions of the effectiveness of various medical interventions from our previous research [28, 29, 30, 31, 32, 33, 34] and adapted query criteria from the review objectives. This dataset enriches our training set on intervention studies and can be used to understand how model generalises to this study type.

#### Algorithm 1

Find the best threshold with the lowest FPR at 100% recall. *value* is the priority score predicted by the model. *label* is the true label of an inclusion/exclusion decision for a study. The algorithm aims to find the threshold with the best performance for the given evaluation metrics.

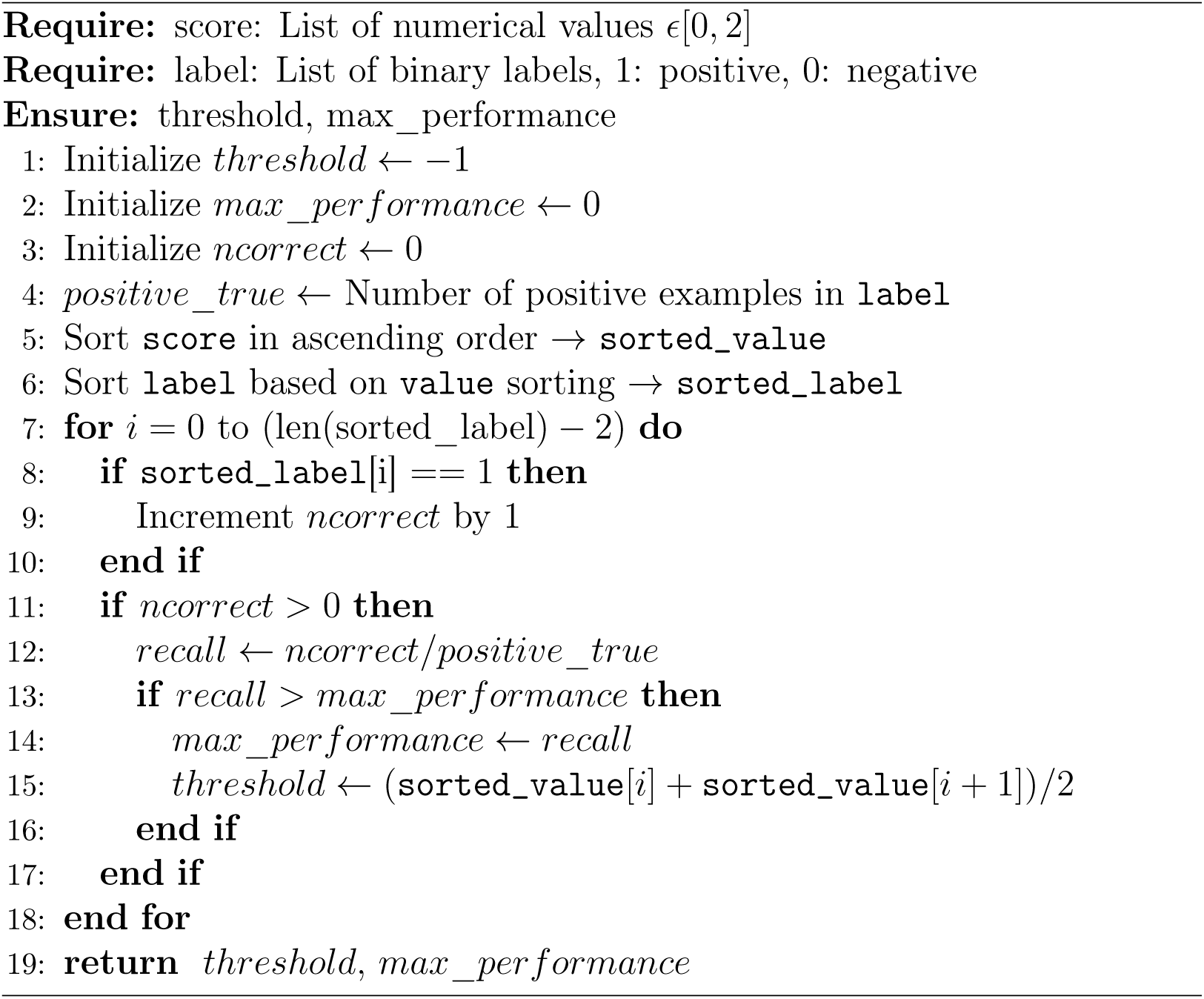

*SYNERGY* [35]. This is an open study screening dataset consisting of 26 systematic reviews. In this study, we removed three reviews for computer science, which resulted in 23 review topics left among medicine, psychology, and biology domains. SYNERGY did not provide selection criteria. Therefore, we collected the selection criteria for each topic from the published systematic review.

The systematic review topics and screening records were grouped into three categories: COVID-19, effectiveness studies (ES), and SYNERGY, where ES includes three non-COVID-19 studies from GUO and all the topics from the Intervention Effectiveness Dataset. The varied heterogeneity within and across these three datasets enables evaluation of how the model performs with more versus less diverse training data. We split each of these three datasets into training (70%), validation (10%) and test (20%) sets, stratifying by review topic. Table 1 shows the basic information for each training set. An overview for all topics included is listed in the supplementary materials (Table B.7).

**Table 1:** Overview of training sets for the general study screening model. Average inclusion percentage is the macro average of the inclusion percentages for all topics in the dataset.

| Dataset | Number of Topics | Number of Inclusion | Number of Records | Average Inclusion Percentage (Minimum, Maximum) |
| --- | --- | --- | --- | --- |
| COVID-19 | 7 | 923 | 27, 169 | 5.05 (0.71, 10.98) |
| ES | 9 | 609 | 26, 473 | 3.54 (0.45, 7.95) |
| SYNERGY | 23 | 1, 842 | 106, 315 | 4.70 (0.16, 21.96) |

### 4.2. Implementation Details

We first trained the model by fine-tuning BlueBERT on the COVID-19 training set and extended the training process with more diverse data from ES and SYNERGY. Four NVIDIA GeForce RTX 2080 Ti GPUs were used to train the model.

During training, we applied a maximum sequence length of 512 tokens (roughly 350–400 words) due to the limitation of BERT, which means a topic or study with more than 512 tokens was truncated. Then, we optimised the model using Online Contrastive Loss and AdamW optimiser [36] with a margin of 0.5 and a learning rate of 2*e ↑* 5. The aim was to increase the distance between irrelevant studies and systematic review topics in the embedding space while decreasing the distance for the relevant studies during training. We trained the model for 6 epochs with a batch size of 6 for each dataset. The model’s hyperparameters, including learning rate, margin, epochs and batch size, were tuned empirically with training and validation sets, and the model was evaluated on the test set.

The binary decision thresholds are calculated on the training set and evaluated on the test set.

### 4.3. Exploring the impact of including selection criteria in topic descriptions

BERT and its derivative models are able to capture the semantic meaning and contextual information. Therefore, adding more contextual information on the topic could potentially improve the study screening performance. To validate this assumption, we added selection criteria to the topic and compared it with the model trained without criteria using the COVID-19 dataset.

Selection criteria are the pre-specified eligibility criteria for determining which studies are included or excluded from the review. They are specified based on the review question and include aspects such as the study participants, settings, treatments of interest, and appropriate study designs. The input for the systematic review topics is structured as follows.

Query: {Systematic review title}. Criteria: {Selection criteria}

We extracted and curated three types of criteria for the COVID-19 dataset: free text criteria, structured criteria, and inclusion/exclusion criteria. Free text criteria are the descriptions of the study selection criteria, which we extracted from the methods section in the published systematic review papers. In the real world however, reviewers tend to have structured criteria that contain information about participants, interventions, comparators, outcomes, and study designs. Thus, we built the structured criteria for all topics in the COVID-19 dataset. In Guo et al.’s study [11], the criteria were defined as a list of inclusion criteria and a list of exclusion criteria. We curated our criteria following the same framework and compared the performance between different types of criteria.

The criteria used for the COVID-19 dataset can be found in supplementary material Table C.8.

### 4.4. Exploring the impact of differences in dataset size and topic homogeneity on predictive performance

We trained the models on the three selected COVID-19 efficiency studies (the same as Guo et al. [11]), all COVID-19 topics, COVID-19 topics with ES topics, and COVID-19 topics with ES and SYNERGY topics, respectively. The trained models were evaluated on all topics in the COVID-19 dataset. By comparing the performance of the study screening models trained with different numbers of topics and topics from various domains, we can understand whether adding more training data and diverse topics can improve the performance on a target theme (COVID-19).

To investigate how the data homogeneity affects the model performance, we calculated a measure of textual similarity between the selected COVID-19 topics and other datasets. Firstly, the topics in each dataset were embedded with Sentence-BERT [26]. The average of the selected topic embeddings in the COVID-19 dataset was considered the centroid of the dataset. Then, we compared the cosine similarity between the centroid and other topics. The similarity between the selected COVID-19 topics and other datasets was calculated as Equation 3.

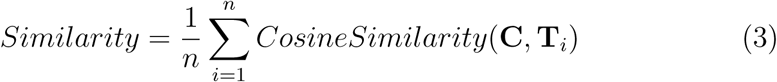

Where *n* is the number of topics in a dataset, **C** is the centroid of the three selected COVID-19 topics, and **T** is the topic embedding.

### 4.5. Evaluating the models performance on unseen topics: A Leave-One-Topic-Out analysis

The model’s ability to generalise to topics not used in training was assessed using the seven topics from the COVID-19 dataset. We used a leave-one-topic-out cross validation approach, where the dataset is split by topic. For each cross-validation iteration, six topics are used to fine-tune BlueBERT and the trained model was evaluated on the remaining topic. If the trained models achieved good performance among target datasets, this would indicate that M-PreSS has a good generalisation of COVID-19 efficacy studies and could be used for new reviews in this area.

### 4.6. Evaluation

We evaluated our models trained with various strategies on the COVID-19 dataset. Table 2 gives an overview of the topics included in the dataset. Seven systematic reviews were included in the dataset; most of them have fewer than 10% of the studies labelled as ‘included’, hence, imbalanced.

**Table 2:** Overview of the COVID-19 dataset. A lower inclusion percentage indicates greater imbalance within the topic.

| Abbreviation | Topic | Inclusion Count | Exclusion Count | Inclusion Percentage |
| --- | --- | --- | --- | --- |
| NMABS | Clinical efficacy and safety of SARS-CoV-2-neutralizing monoclonal antibody in patients with COVID-19: A living systematic review and meta-analysis | 297 | 2, 801 | 9.59% |
| CSR | Efficacy and safety of corticosteroid regimens for the treatment of hospitalized COVID-19 patients: a meta-analysis | 140 | 8, 865 | 1.55% |
| IVM | Efficacy and safety of ivermectin for the treatment of COVID-19: a systematic review and meta-analysis | 35 | 279 | 11.15% |
| SSRI | Efficacy and safety of selective serotonin reuptake inhibitors in COVID-19 management: a systematic review and meta-analysis | 29 | 3, 989 | 0.72% |
| CQHCQ | Efficacy of chloroquine and hydroxychloroquine for the treatment of hospitalized COVID-19 patients: a meta-analysis | 311 | 8, 148 | 3.68% |
| LPVR | Efficacy of lopinavir-ritonavir combination therapy for the treatment of hospitalized COVID-19 patients: a meta-analysis | 91 | 1, 456 | 5.88% |
| PMH | Prevalence of mental health symptoms in children and adolescents during the COVID-19 pandemic: A meta-analysis | 422 | 13, 280 | 3.08% |

Due to the imbalance of the dataset, using accuracy metric can be misleading because it does not sufficiently reflect the model’s effectiveness across all classes. In our datasets, where the number of included studies significantly outnumbers the exclusions, a high accuracy score can be achieved by predicting the exclusion class for all instances. Therefore, we evaluated different trained study screening models with PRAUC (Area under the Precision-Recall Curve), which focuses on the performance relative to the inclusion (minority) class. A perfect prediction would have a PRAUC equal to 1. The PRAUC of a random model would equal the inclusion percentage, meaning that a model with a PRAUC greater than the inclusion percentage can discriminate between inclusion and exclusion to some extent.

To further evaluate the performance of our model, we compared our trained COVID-19 study screening models with the results reported by Guo et al. In Guo et al.’s study, the GPT model was required to give a decision on inclusion/exclusion for a candidate study. Therefore, we applied a binary threshold to the COVID-19 model for comparison. The thresholds were calculated respectively for each review topic using the best FPR at 100% recall strategy (explained in Section 3.1).

When evaluating the performance of inclusion/exclusion prediction, we calculated the recall (equation 1), FPR (equation 2), and true negative rate (TNR, also known as specificity) as follows.

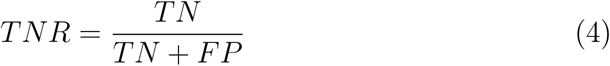

Recall indicates how well the model captured all the included studies, which is a key metric for study screening. FPR is the percentage of exclusion examples predicted as inclusion by the model, which indicates the extra irrelevant studies needed to review for the reviewers. TNR (also known as specificity) indicates how well the model correctly identifies exclusion examples from all actual excluded studies.

## 5. Results and Discussions

### 5.1. Exploring the impact of including selection criteria in topic descriptions

Figure 2 compares the performance of the study screening models trained with and without selection criteria. As shown in the figure, the PRAUC of all the models is better than the baseline, which is a random classifier. Furthermore, the model trained with criteria achieved a comparable or better performance than the model without them, for all except one topic (CSR). This shows that extra contextual information on selection criteria provided to the model is beneficial for distinguishing included references from excluded ones. Therefore, criteria were added to the inputs during training for all the models in this study.

Our model trained with selection criteria (free text) achieved an average PRAUC of 0.61 among the COVID-19 review topics and had the best performance of 0.84 PRAUC on IVM. Furthermore, the PRAUC among all topics was higher than the inclusion percentages, which shows that our model is better than a random classifier and can discriminate between inclusion and exclusion studies.

One limitation of using BlueBERT is that the input sequence length has a limitation of 512 tokens, typically representing around 350–400 words. For any input that is more than 512 tokens, the model will truncate the input text. Therefore, using Siamese architecture can separate the selection criteria and study title and abstract into two identical language models, hence minimising the loss of information from the review topics and studies.

When pre-training the study selection model with a larger number of topics, the free text criteria are more accessible from the published paper than manually curated criteria. However, to further understand the role of selection criteria, we compared the free text criteria with other curated criteria.

The results, provided in Table 3, show that the performance of different criteria varies between topics. Free text achieved the highest PRAUC among four out of seven topics, followed by structured criteria (three out of seven) and inclusion/exclusion criteria (two out of seven). The structured and inclusion/exclusion criteria had a comparable or higher recall for all topics than free text. Structured criteria achieved better recall on SSRI, LPVR, and PMH, whereas inclusion/exclusion criteria outperformed structured criteria on NMBAS, CSR, and CQHCQ. Nevertheless, the structured criteria performed better in terms of average recall and average FPR, which shows that the model (with our choice of classification threshold) is better at including all the relevant studies while adding fewer irrelevant ones.

**Table 3:** Performance of the models trained with different formats of selection criteria. The criteria for each topic can be found at Appendix C. Free text criteria are the study selection description extracted from the published systematic review. Structured criteria are the curated selection objectives, which include participants, interventions, comparators, outcomes, and study designs. Inclusion/Exclusion criteria are the criteria curated in an inclusion and exclusion format.

| Topic | Free Text |  |  | Structured |  |  | Inclusion/Exclusion |  |  |
| --- | --- | --- | --- | --- | --- | --- | --- | --- | --- |
|  | PRAUC | Recall | FPR(%) | PRAUC | Recall | FPR(%) | PRAUC | Recall | FPR(%) |
| NMABS | <u>0.80</u> | 0.93 | <u>10.16</u> | 0.74 | 0.95 | 11.41 | 0.76 | <u>0.97</u> | 13.55 |
| CSR | 0.53 | 0.86 | <u>1.24</u> | <u>0.77</u> | 0.86 | 2.31 | 0.53 | <u>0.93</u> | 6.77 |
| IVM | <u>0.84</u> | <u>1.00</u> | 19.64 | 0.59 | <u>1.00</u> | 7.14 | 0.80 | <u>1.00</u> | <u>3.57</u> |
| SSRI | 0.29 | <u>0.67</u> | 0.88 | 0.33 | <u>0.67</u> | 1.38 | <u>0.46</u> | 0.50 | <u>0.63</u> |
| CQHCQ | <u>0.60</u> | 0.79 | <u>5.40</u> | <u>0.60</u> | 0.84 | 5.58 | 0.51 | <u>0.86</u> | 8.65 |
| LPVR | <u>0.48</u> | 0.63 | <u>5.82</u> | 0.45 | <u>0.74</u> | 10.62 | 0.47 | 0.68 | 7.53 |
| PMH | 0.69 | 0.89 | <u>6.40</u> | <u>0.70</u> | <u>0.95</u> | 6.93 | <u>0.70</u> | 0.91 | <u>4.97</u> |
| **Average** | <u>0.61</u> | 0.83 | 7.08 | 0.60 | <u>0.86</u> | <u>6.48</u> | 0.60 | 0.83 | 6.52 |

In Table 4, we compared the model trained with inclusion/exclusion criteria with Guo et al.’s results using ChatGPT/GPT-4. The thresholds that achieved the lowest FPR at 100% recall were applied to obtain inclusion/exclusion predictions. Note that Guo et al. evaluated the GPT model on the entire study screening dataset without training or fine-tuning the model. However, by fine-tuning BlueBERT with 80% screening records from the COVID-19 dataset, we evaluated our model only on the sampled test data from the original dataset.

**Table 4:** Model performance compared to Guo et al. [11]. We trained the COVID-19 model with the training set of all seven topics from the COVID-19 datasets. The inclusion/exclusion selection criteria were included in the input during the training. A threshold for each topic was calculated to achieve the lowest FPR at 100% recall in the training set. Note that Guo et al. evaluated the performance with all the records in each topic. However, we trained our model with 80% of the records. Hence, our models were evaluated on the test set (20% of the records) for each topic.

| Topic | Guo et al. |  |  | Ours: COVID-19 Model (best FPR at 100% recall) |  |  |
| --- | --- | --- | --- | --- | --- | --- |
|  | Recall | TNR(%) | FPR (%) | Recall | TNR(%) | FPR(%) |
| IVM | 0.69 | 75.63 | 24.37 | <u>1.00</u> | 96.43 | <u>3.57</u> |
| SSRI | <u>0.97</u> | 94.91 | 5.09 | 0.50 | <u>99.37</u> | <u>0.63</u> |
| LPVR | 0.59 | 86.20 | 13.80 | <u>0.68</u> | <u>92.47</u> | <u>7.53</u> |

Although BlueBERT is a smaller language model compared to the GPT models, the results in Table 4 show that fine-tuning BlueBERT with study screening data can obtain comparable results to a large general model, such as ChatGPT, on the three topics reported in Guo et al.’s study. For IVM and LPVR, our model managed to identify more relevant studies without including additional false positive studies compared to Guo et al.’s results from GPT-4. Moreover, our model achieved 100% recall on IVM, which means it managed to identify all the relevant studies from the candidates.

### 5.2. Exploring the impact of differences in dataset size and topic homogeneity on predictive performance

The evaluation results of training on different datasets, with different dataset sizes and topic homogeneity, are illustrated in Figure 3. To maintain consistency between datasets, we applied free text criteria for all the topics in this experiment. The models were trained with training sets from specific topics and evaluated on the test sets from the COVID-19 dataset. Following Guo et al.’s study, the selected COVID-19 model was trained on IVM, SSRI, and LPVR. Thus, we did not evaluate the selected COVID-19 topics model on other unseen topics.

**Figure 3:**
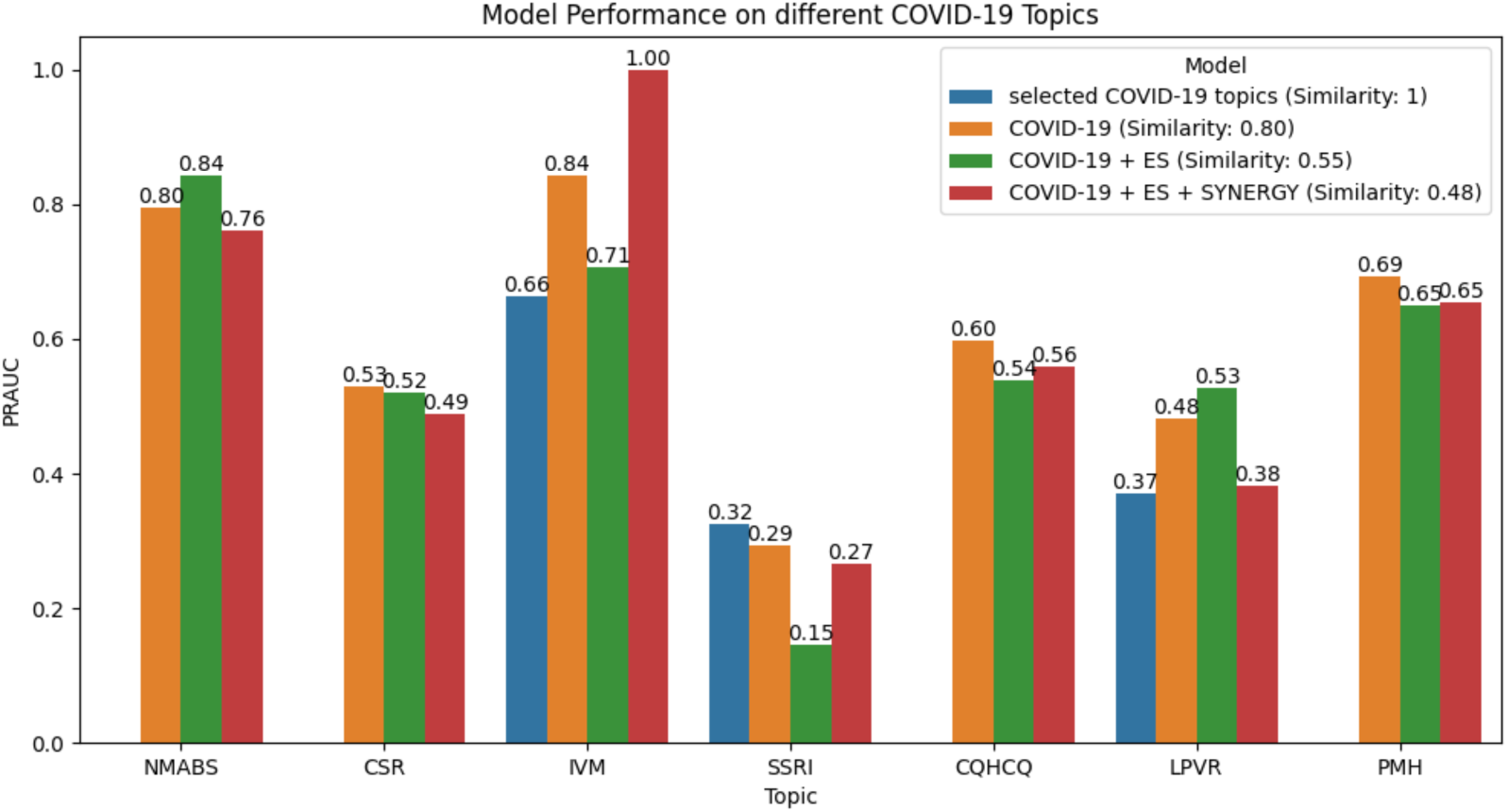
Comparing the performance of models trained on various data sizes (see Table 1). The models were trained on the COVID-19 training set and evaluated on the test set. We measured the average cosine similarity between the centroid of the three selected COVID-19 topics and other topics for training the models (explained in Section 4.4). The selected COVID-19 topics model was only trained on IVM, SSRI, AND LPVR. Therefore, we only evaluated it on these three topics.

The results in Figure show that the model (orange) trained with all COVID-19 topics outperformed the model (blue) trained with selected topics on the IVM and LPVR test set, showing that adding in domain data can improve the model performance. However, the model performance decreased on SSRI when more COVID-19 topics were added during training. This may be because that free text criteria resulted in lower performance than other criteria formats for SSRI but performs better for IVM and LPVR (Table 3). We quantified the data homogeneity by calculating the cosine similarity between the selected COVID-19 and added topics. The similarity dropped from 0.8 to 0.55 and 0.48 when adding ES and SYNERGY datasets. The improvements were inconsistent among all the topics when increasing the size of study screening data from a broader but related domain (e.g. ES and SYNERGY dataset), indicating that the homogeneity of the data matters during scaling.

Furthermore, we compared our model trained on three selected COVID-19 topics with the results from Guo et al. using the same criteria. Similar to the COVID-19 model, we used the best FPR at 100% recall threshold to give a binary prediction for each candidate study. With limited topics, our model still improved the recall for IVM and LPVR (shown in Table 5), demonstrating that fine-tuning with study screening datasets is an effective approach for researchers with constrained computational resources to improve the performance of general LLMs.

**Table 5:** Performance of the model trained with three selected COVID-19 topics compared to the results reported by Guo et al. [11]. We used the same topics and inclusion/exclusion criteria as Guo et al. for training the model.

| Topic | Guo et al. |  |  | Selected COVID-19 Model (best FPR at 100% recall) |  |  |
| --- | --- | --- | --- | --- | --- | --- |
|  | Recall | TNR(%) | FPR (%) | Recall | TNR(%) | FPR(%) |
| IVM | 0.69 | 75.63 | 24.37 | <u>1.00</u> | <u>92.86</u> | <u>7.14</u> |
| SSRI | <u>0.97</u> | 94.91 | 5.09 | 0.17 | <u>99.12</u> | <u>0.88</u> |
| LPVR | 0.59 | 86.20 | 13.80 | <u>0.79</u> | <u>86.64</u> | <u>13.36</u> |

### 5.3. Evaluating the models performance on unseen topics: A Leave-One-Topic-Out analysis

The results so far demonstrated in this study show that our model has a good generalisation among different topics, for the set of topics used in model training. Another advantage of utilising an LLM is that the model performance can potentially be generalised and transferred to unseen data or topics after training. Therefore, we performed a leave-one-topic-out cross validation for all COVID-19 topics to evaluate the transferability of knowledge learned from given study screening records to a new topic. The results are illustrated in figure 4.

**Figure 4:**
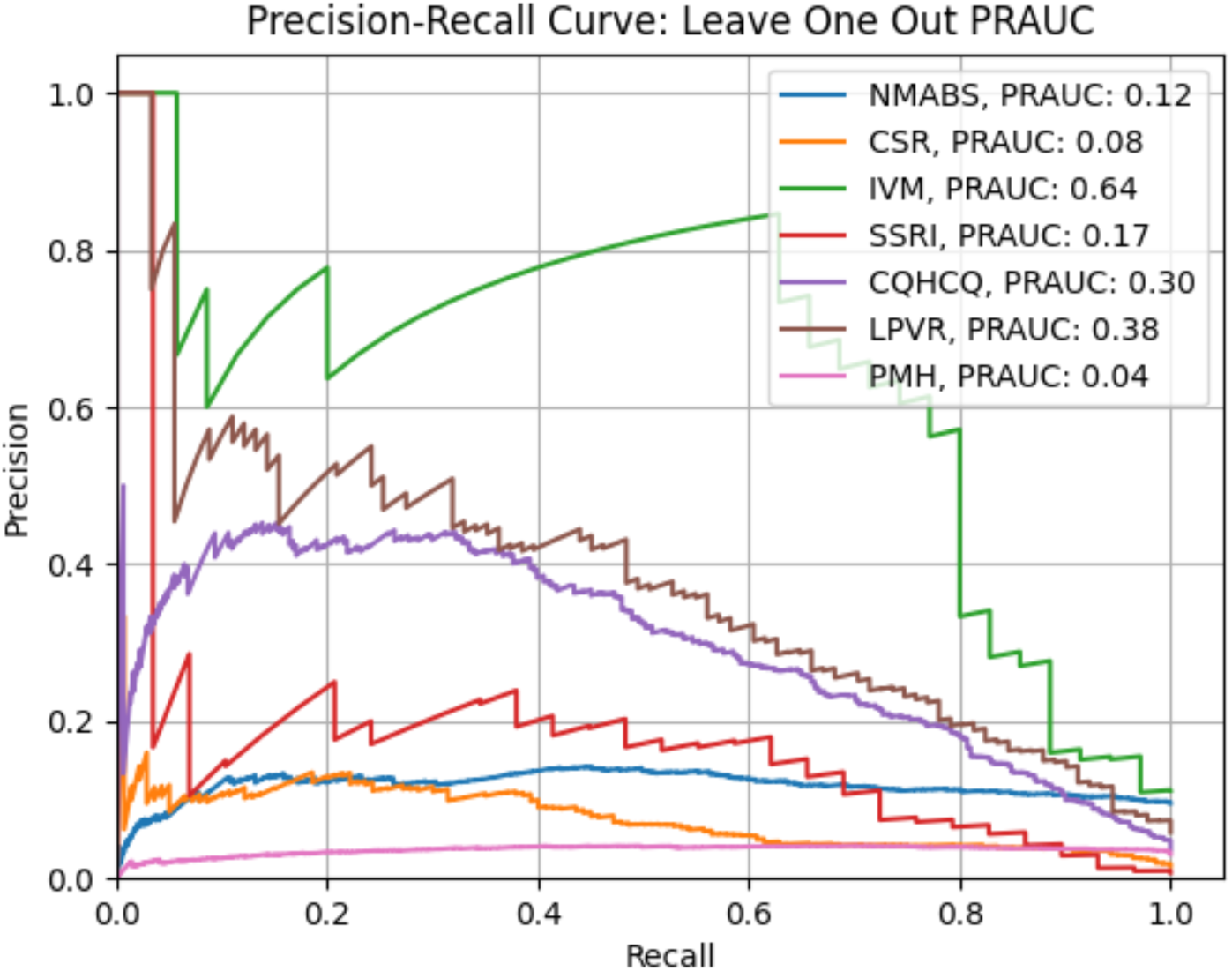
Precision-Recall curves for leave-one-topic-out validation. The performance on each topic is based on zero-shot learning.

The results show that the model performance on unseen topics decreased compared to the results trained with the entire COVID-19 dataset (see free text results in Table 3), as predicting without showing the model examples from the target topic was more challenging. However, the model still discriminated included studies from exclusions to some extent, achieving a larger PRAUC than the topic baseline (i.e. inclusion percentage - see Table 2).

Performance of our model decreased least for LPVR (PRAUC 0.38 vs 0.48) and IVM (PRAUC 0.64 vs 0.84), better than PMH (PRAUC 0.04 vs 0.69) which had the largest decrease. This may be because these LPVR and IVM contain the least number of records, such that removing these examples from the training data does not dramatically affect the model’s performance.

## 6. Conclusion

In this study, we applied a Siamese architecture to fine-tune BlueBERT for study screening. Our results show that our model has good generalisation across different topics, where examples from these topics are used in training. This training method can also be applied to other large language model frameworks, such as Llama ^1^. The models we trained achieved better recall and lower FPR compared to Guo et al.’s study using ChatGPT on two out of three COVID-19 topics. This shows that even with limited data and a smaller language model, fine-tuning can achieve comparable or better performance. We experimented with various formats of selection criteria and found that adding selection criteria during training can increase prediction performance, and the selection criteria curated in the format of participants, interventions, comparators, outcomes, and study designs achieved the best average recall and FPR among the COVID-19 dataset. We also observed that adding more systematic review topics with high homogeneity to the model can potentially increase the overall performance across topics.

Our method shows potential in performing study screening for systematic review topics where the topics are related to a research focus. For example, we applied our method to automate the screening of Global Cancer Update Programme (CUP Global) systematic reviews about cancer incidence topics for a variety of cancer sites [37], and achieved good results where the recall for all topics are consistently above 90% while the FPRs are under 10% for the majority of the topics. Our developed model can also be used for other study screening processes in a similar setting. In addition, this method could be applied in a more semi-automated manner to assist expert decision-making, where the developed model can be used under different thresholds to achieve multiple inclusion/exclusion strategies (such as identifying definitely included and excluded studies), or being combined with more rule-based approaches for weighted scoring.

However, transferring the screening knowledge learned from existing topics to new topics with zero-shot learning remains challenging for our model powered by BlueBERT. In future research, few-shot learning (labels a small amount of data) or active learning could be applied to fine-tune the pretrained study screening model for better performance on new review topics. For instance, with active learning, human reviewers can review the top K studies prioritised by the pre-trained model. Then, the decisions could be fed back to tuning the study screening model to improve the accuracy. This process can be repeated until the model achieves a better prediction.

## Declarations

### Data and Code Availability

The code will be available on GitHub^2^when published. The open dataset can be found at:

- Guo et al.: https://data.mendeley.com/datasets/np79tmhkh5/1
- SYNERGY: https://github.com/asreview/synergy-dataset

## Data Availability

All data produced in the present study are available upon reasonable request to the authors.

## Acknowledgements

This study is part of a research project on the automation of systematic review processes in the Global Cancer Update Programme (CUP Global). CUP Global produced by World Cancer Research Fund (WCRF) International, is a rigorous systematic research programme to analyse and judge global research on how diet, nutrition, physical activity and body weight affect cancer risk and survival. Findings and evidence judgements are used to develop public health recommendations for cancer prevention and recommendations for future research.

## Appendix A. Data examples for M-PreSS

Table A.6 gives example inputs and outputs for the M-PreSS framework.

**Table A.6:** Input and Output Examples for M-PreSS.

| Systematic Review Topic | Candidate Study | Priority Score |
| --- | --- | --- |
| Query: Prevalence of mental health symptoms in children and adolescents during the COVID-19 pandemic: A meta-analysis | Title: COVID-19 and Adolescent Depression and Suicide Risk Screening Outcomes Abstract: BACKGROUND: Mental health concerns increased during the coronavirus disease 2019 pandemic, but previous studies have not examined depression screening in pediatric primary care. We aimed to describe changes in screening, depressive symptoms, and suicide risk among adolescents during the coronavirus disease 2019 pandemic. METHODS: In a repeat cross-sectional analysis of electronic health record data from a large pediatric primary care network, we compared the percentage of primary care visits where adolescents aged 12 to 21 were screened for depression, screened positive for depressive symptoms, or screened positive for suicide risk between June and December 2019 (prepandemic) and June and December 2020 (pandemic). Changes were examined overall, by month, and by sex, race and ethnicity, insurance type, and income. Modified Poisson regression was used to calculate prevalence ratios (PRs) for the prepandemic to pandemic changes. RESULTS: Depression screening at primary care visits declined from 77.6% to 75.8% during the pandemic period (PR: 0.98, 95% confidence interval [CI]: 0.90-1.06). The percentage of adolescents screening positive for depressive symptoms increased from 5.0% to 6.2% (PR: 1.24, 95% CI: 1.15-1.34), with greater increases among female, non-Hispanic Black, and non-Hispanic white adolescents. Positive suicide risk screens increased from 6.1% to 7.1% (PR: 1.16, 95% CI: 1.08-1.26), with a 34% relative increase in reporting recent suicidal thoughts among female adolescents (PR: 1.34, 95% CI: 1.18-1.52). CONCLUSIONS: Results suggest that depression and suicide concerns have increased during the pandemic, especially among female adolescents. Results underscore the importance of consistent depression and suicidality screening. | 0.4994128 |

| **Systematic Review Topic** | **Candidate Study** | **Priority Score** |
| --- | --- | --- |
| Query: Prevalence of mental health symptoms in children and adolescents during the COVID-19 pandemic: A meta-analysis | Title: Hopes and fears: emerging adults in mood and anxiety disorder treatment predict outcomes of the COVID-19 pandemic Abstract: Active patients at The First Episode Mood and Anxiety Program (FEMAP) were asked to participate in a study monitoring their symptoms during early COVID-19 closures. Participants were asked to predict both possible positive and feared outcomes of the pandemic. Responses were analyzed with a descriptive qualitative approach using thematic analysis. Primary feared outcomes included death, prolonged pandemic impacts, and concerns about employment, mental health, and education. Prevalent positive predictions included a new perspective, improved relationships, personal growth, and environmental improvement. These findings suggest the complex relationship between fear and hope for young people in the early stages of the pandemic. | 0.4899408 |

## Appendix B. Topic overview for training M-Press

Table B.7 gives an overview of the systematic review topics used in this study.

**Table B.7:**
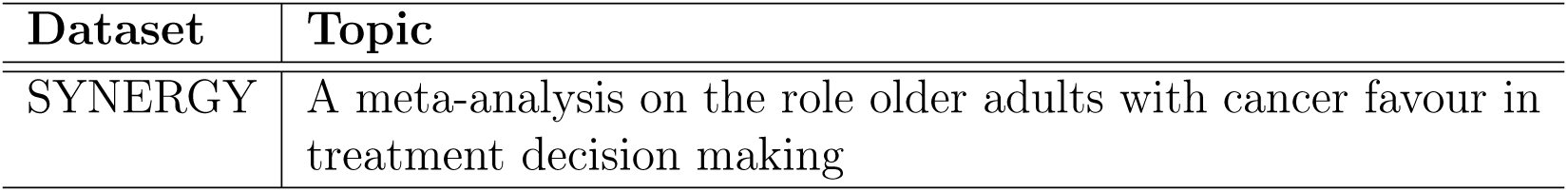
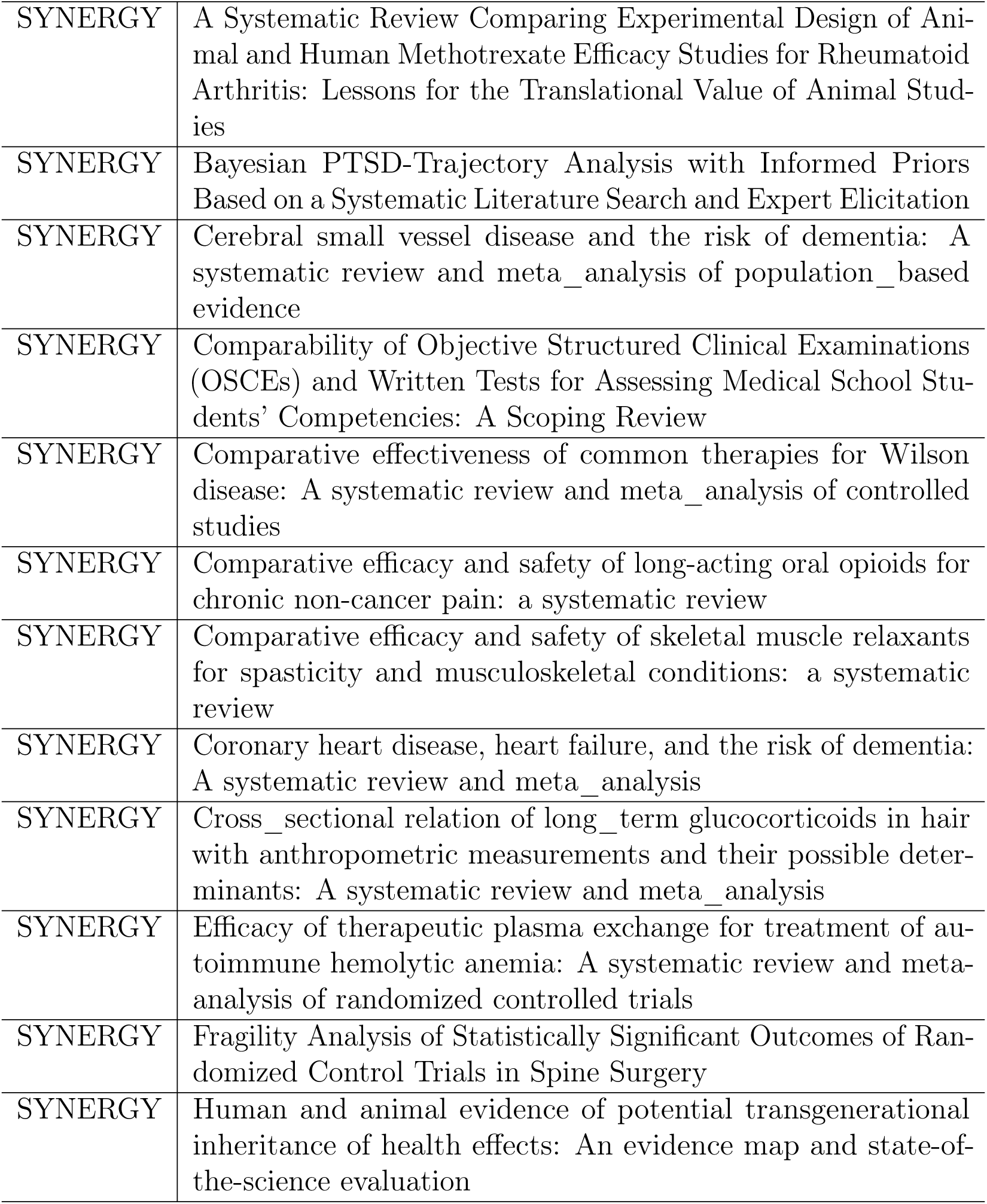
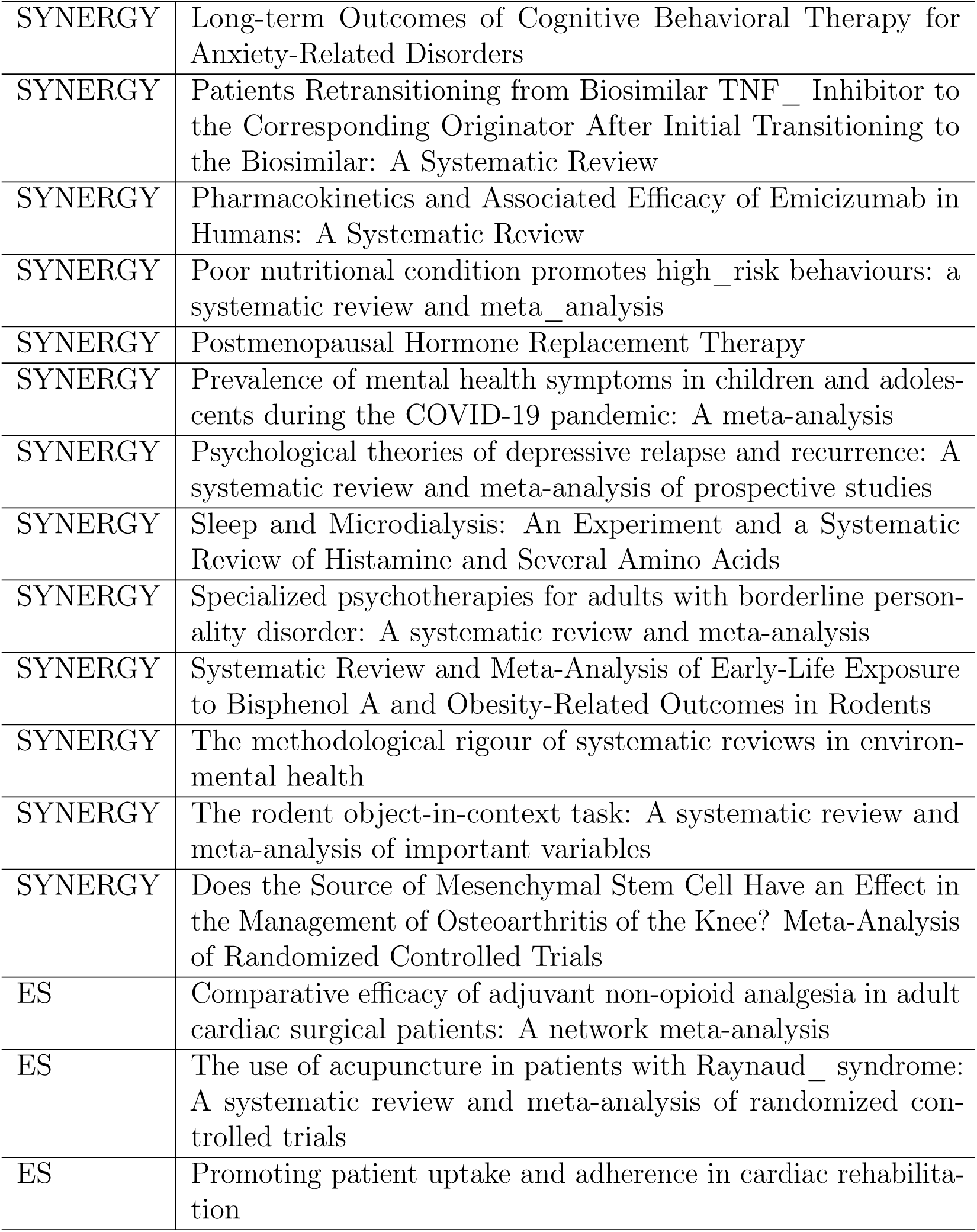
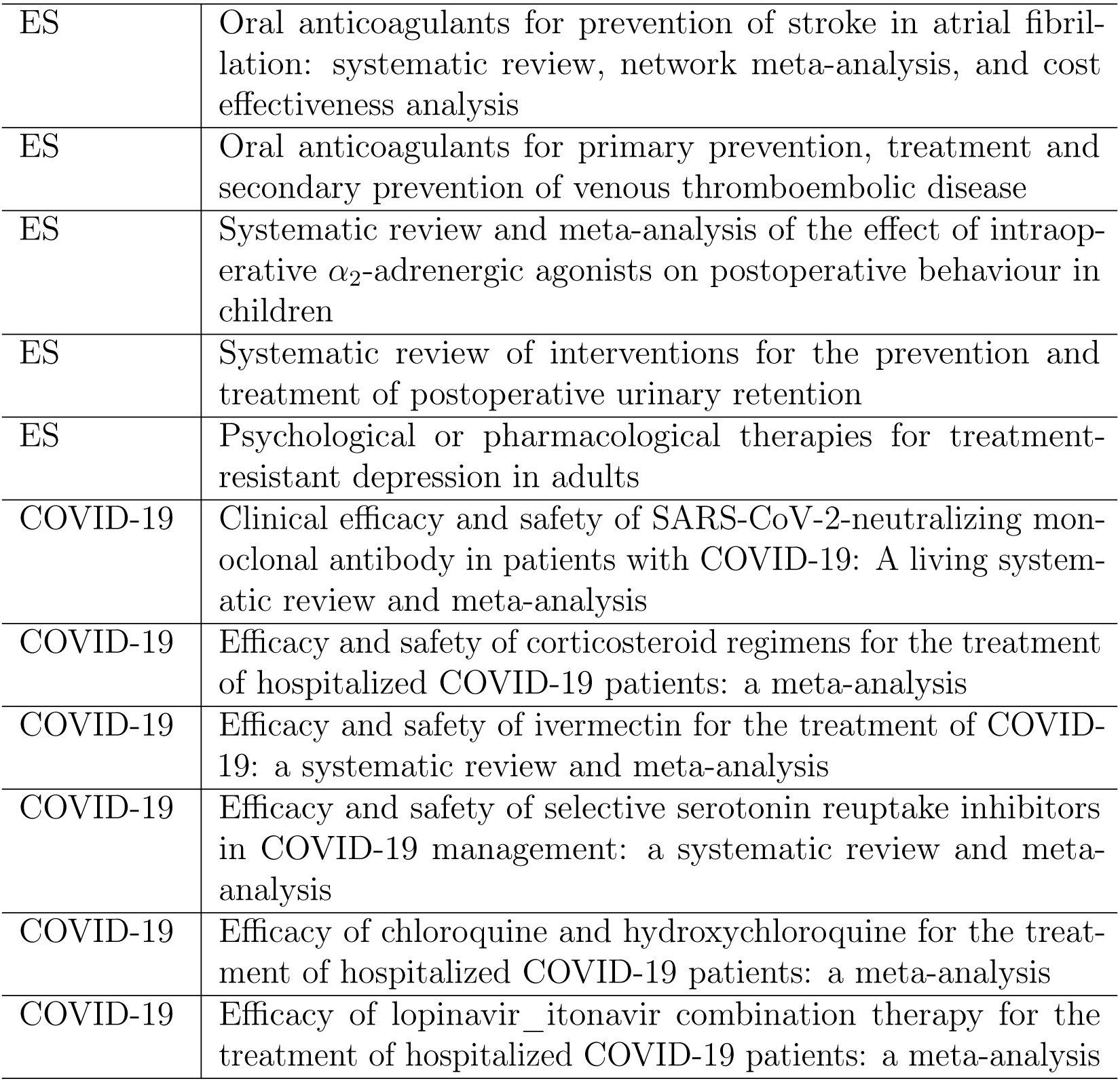
Topics Overview. Topics used for training M-PreSS.

| **Dataset** | **Topic** |
| --- | --- |
| SYNERGY | A meta-analysis on the role older adults with cancer favour in treatment decision making |
| SYNERGY | A Systematic Review Comparing Experimental Design of Animal and Human Methotrexate Efficacy Studies for Rheumatoid Arthritis: Lessons for the Translational Value of Animal Studies |
| SYNERGY | Bayesian PTSD-Trajectory Analysis with Informed Priors Based on a Systematic Literature Search and Expert Elicitation |
| SYNERGY | Cerebral small vessel disease and the risk of dementia: A systematic review and meta_analysis of population_based evidence |
| SYNERGY | Comparability of Objective Structured Clinical Examinations (OSCEs) and Written Tests for Assessing Medical School Students' Competencies: A Scoping Review |
| SYNERGY | Comparative effectiveness of common therapies for Wilson disease: A systematic review and meta_analysis of controlled studies |
| SYNERGY | Comparative efficacy and safety of long-acting oral opioids for chronic non-cancer pain: a systematic review |
| SYNERGY | Comparative efficacy and safety of skeletal muscle relaxants for spasticity and musculoskeletal conditions: a systematic review |
| SYNERGY | Coronary heart disease, heart failure, and the risk of dementia: A systematic review and meta_analysis |
| SYNERGY | Cross_sectional relation of long_term glucocorticoids in hair with anthropometric measurements and their possible determinants: A systematic review and meta_analysis |
| SYNERGY | Efficacy of therapeutic plasma exchange for treatment of autoimmune hemolytic anemia: A systematic review and meta-analysis of randomized controlled trials |
| SYNERGY | Fragility Analysis of Statistically Significant Outcomes of Randomized Control Trials in Spine Surgery |
| SYNERGY | Human and animal evidence of potential transgenerational inheritance of health effects: An evidence map and state-of-the-science evaluation |
| SYNERGY | Long-term Outcomes of Cognitive Behavioral Therapy for Anxiety-Related Disorders |
| SYNERGY | Patients Retransitioning from Biosimilar TNF_ Inhibitor to the Corresponding Originator After Initial Transitioning to the Biosimilar: A Systematic Review |
| SYNERGY | Pharmacokinetics and Associated Efficacy of Emicizumab in Humans: A Systematic Review |
| SYNERGY | Poor nutritional condition promotes high_risk behaviours: a systematic review and meta_analysis |
| SYNERGY | Postmenopausal Hormone Replacement Therapy |
| SYNERGY | Prevalence of mental health symptoms in children and adolescents during the COVID-19 pandemic: A meta-analysis |
| SYNERGY | Psychological theories of depressive relapse and recurrence: A systematic review and meta-analysis of prospective studies |
| SYNERGY | Sleep and Microdialysis: An Experiment and a Systematic Review of Histamine and Several Amino Acids |
| SYNERGY | Specialized psychotherapies for adults with borderline personality disorder: A systematic review and meta-analysis |
| SYNERGY | Systematic Review and Meta-Analysis of Early-Life Exposure to Bisphenol A and Obesity-Related Outcomes in Rodents |
| SYNERGY | The methodological rigour of systematic reviews in environmental health |
| SYNERGY | The rodent object-in-context task: A systematic review and meta-analysis of important variables |
| SYNERGY | Does the Source of Mesenchymal Stem Cell Have an Effect in the Management of Osteoarthritis of the Knee? Meta-Analysis of Randomized Controlled Trials |
| ES | Comparative efficacy of adjuvant non-opioid analgesia in adult cardiac surgical patients: A network meta-analysis |
| ES | The use of acupuncture in patients with Raynaud_ syndrome: A systematic review and meta-analysis of randomized controlled trials |
| ES | Promoting patient uptake and adherence in cardiac rehabilitation |
| ES | Oral anticoagulants for prevention of stroke in atrial fibrillation: systematic review, network meta-analysis, and cost effectiveness analysis |
| ES | Oral anticoagulants for primary prevention, treatment and secondary prevention of venous thromboembolic disease |
| ES | Systematic review and meta-analysis of the effect of intraoperative $\alpha_2$-adrenergic agonists on postoperative behaviour in children |
| ES | Systematic review of interventions for the prevention and treatment of postoperative urinary retention |
| ES | Psychological or pharmacological therapies for treatment-resistant depression in adults |
| COVID-19 | Clinical efficacy and safety of SARS-CoV-2-neutralizing monoclonal antibody in patients with COVID-19: A living systematic review and meta-analysis |
| COVID-19 | Efficacy and safety of corticosteroid regimens for the treatment of hospitalized COVID-19 patients: a meta-analysis |
| COVID-19 | Efficacy and safety of ivermectin for the treatment of COVID-19: a systematic review and meta-analysis |
| COVID-19 | Efficacy and safety of selective serotonin reuptake inhibitors in COVID-19 management: a systematic review and meta-analysis |
| COVID-19 | Efficacy of chloroquine and hydroxychloroquine for the treatment of hospitalized COVID-19 patients: a meta-analysis |
| COVID-19 | Efficacy of lopinavir_ritonavir combination therapy for the treatment of hospitalized COVID-19 patients: a meta-analysis |

## Appendix C. Selection criteria for COVID-19 topics

Table C.8 list all criteria used when training the COVID-19 systematic review topics. Three types criteria were included: free text, structured, and inclusion/exclusion criteria.

**Table C.8:**
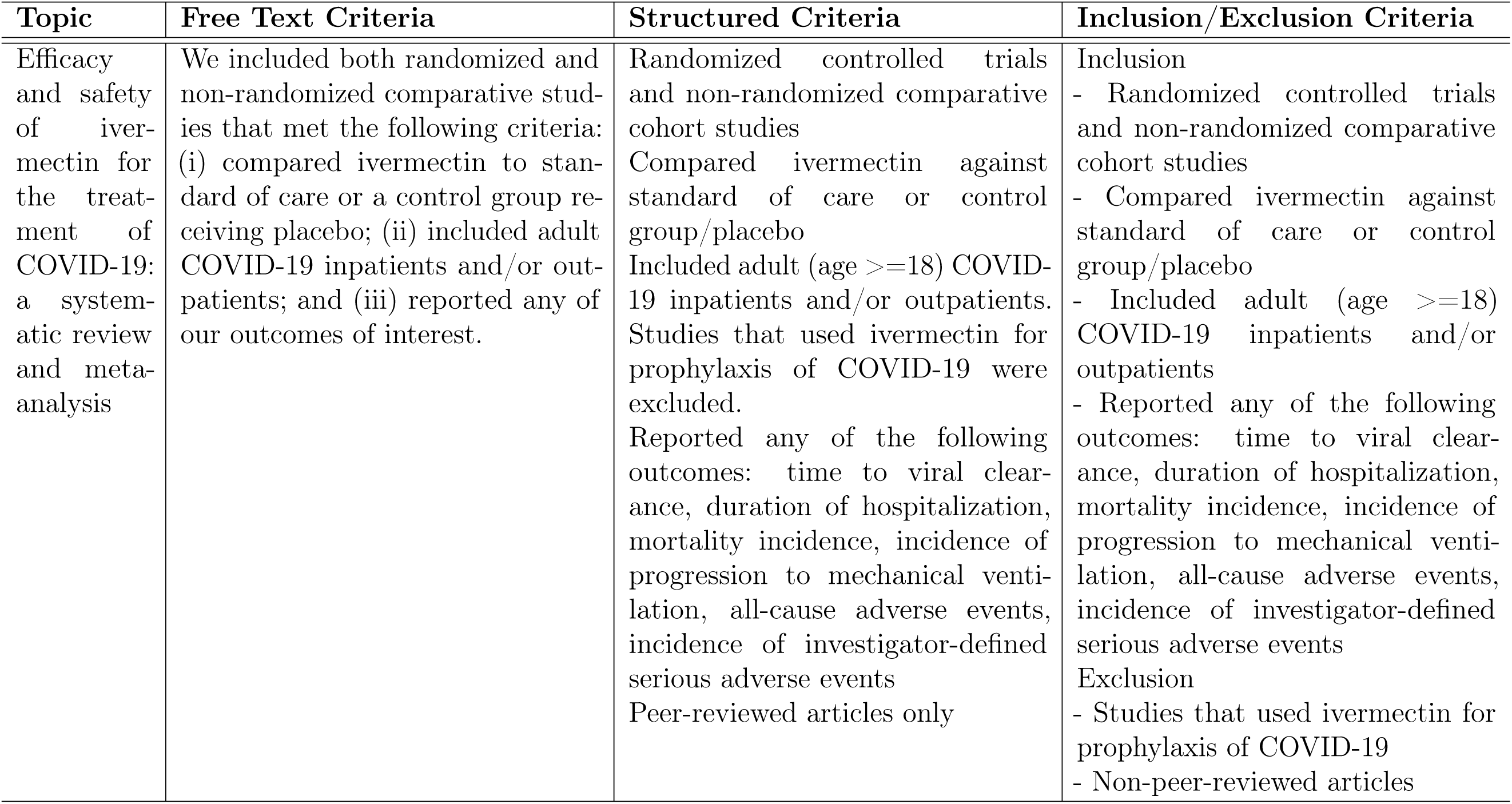
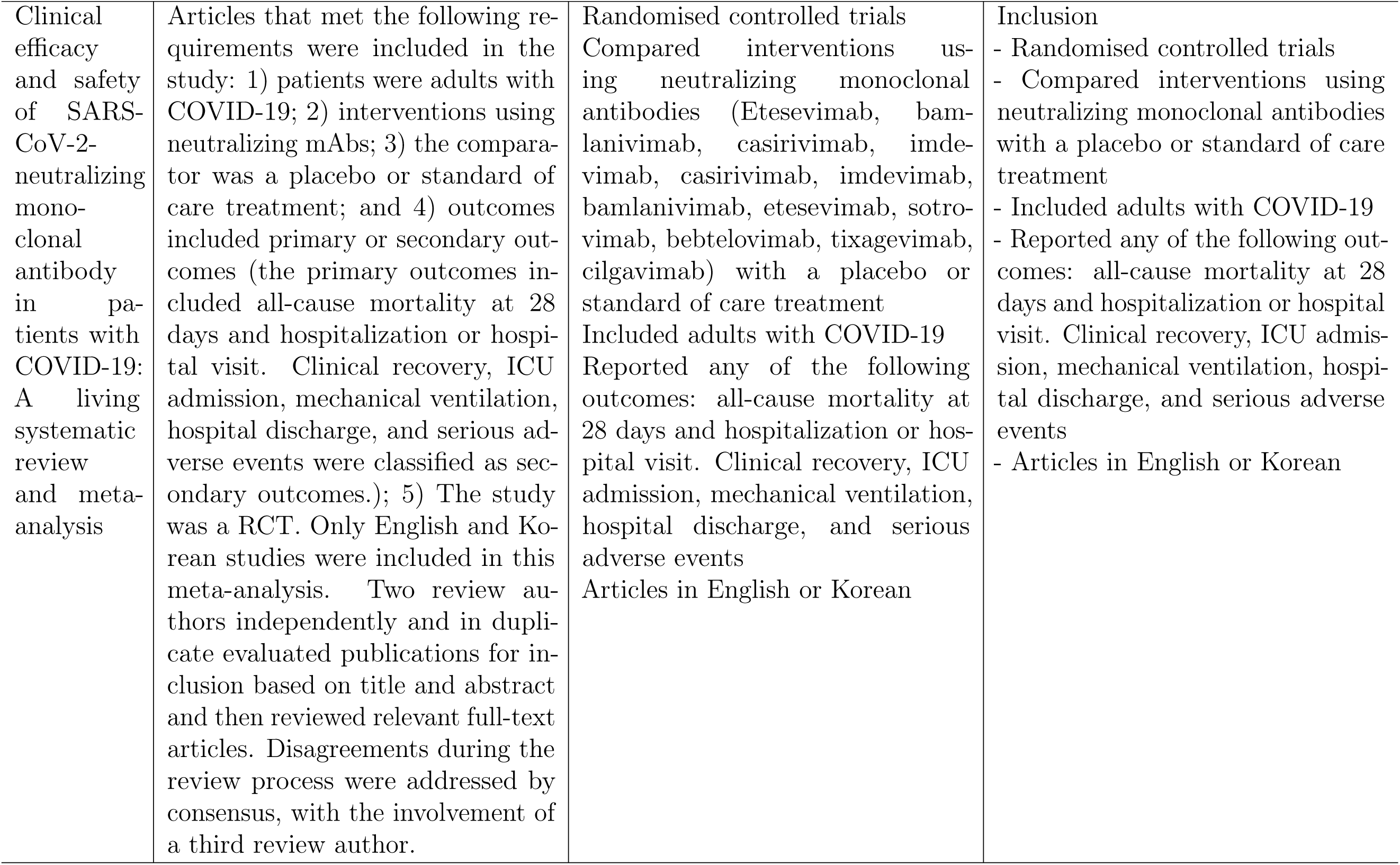
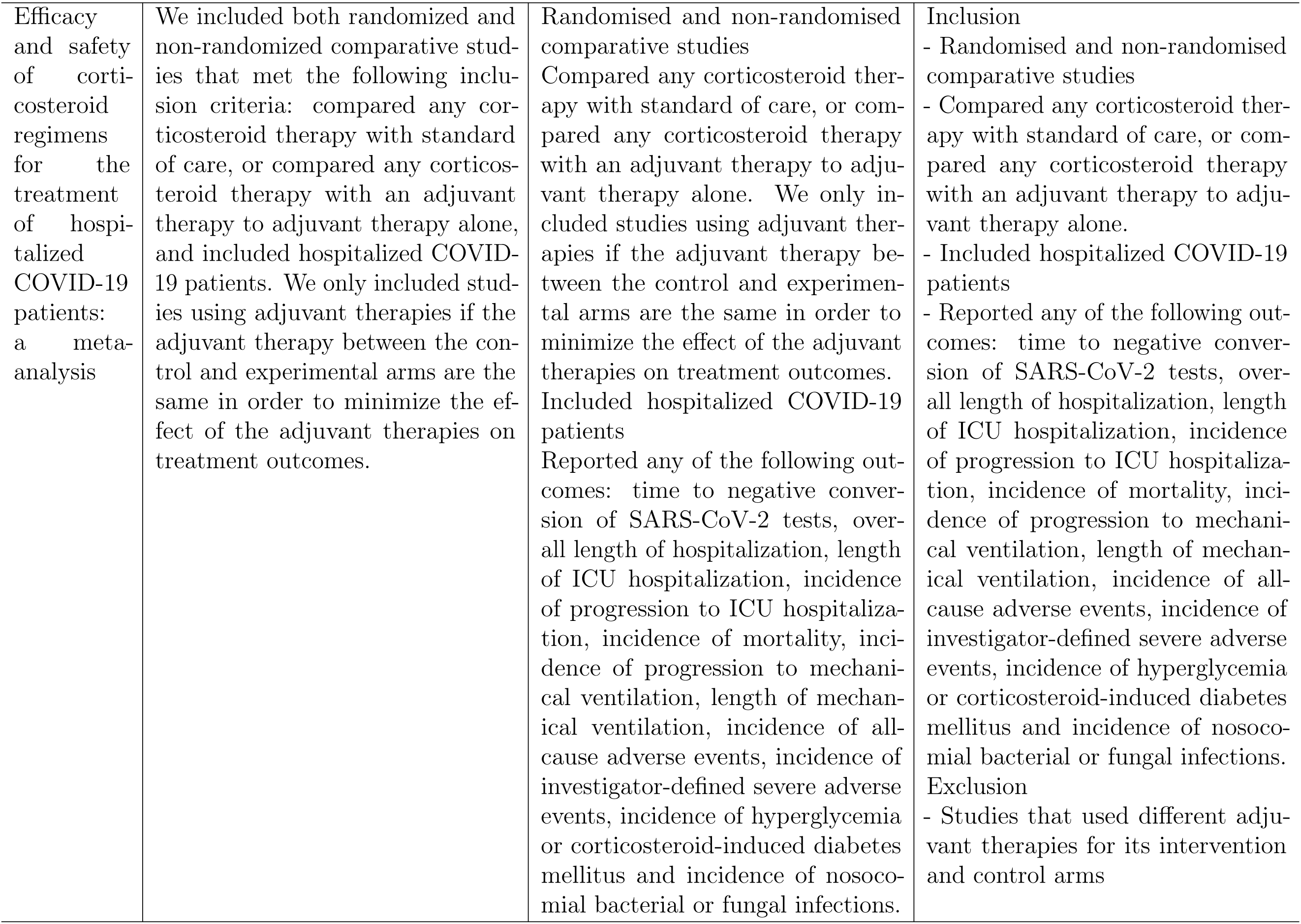
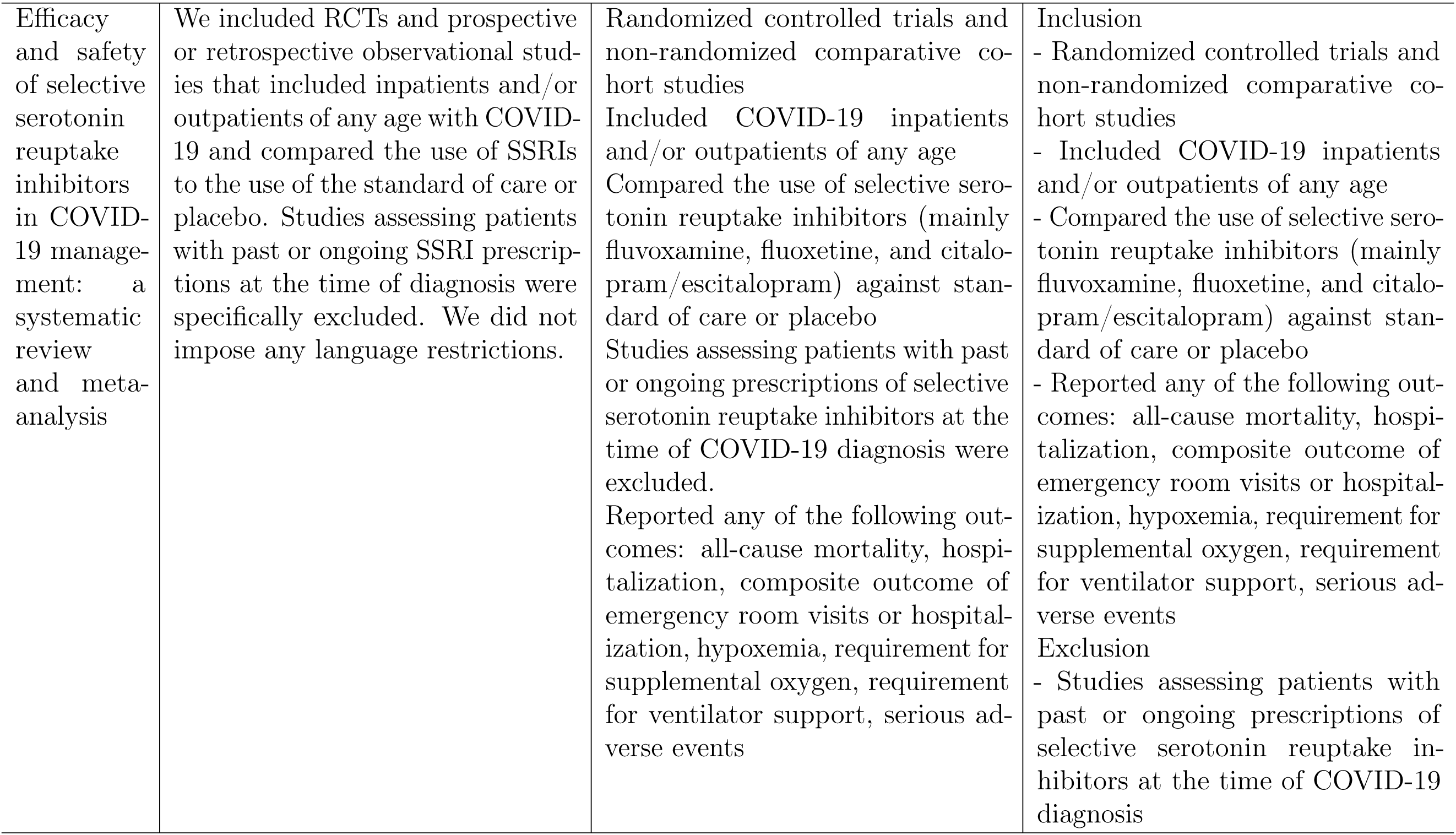
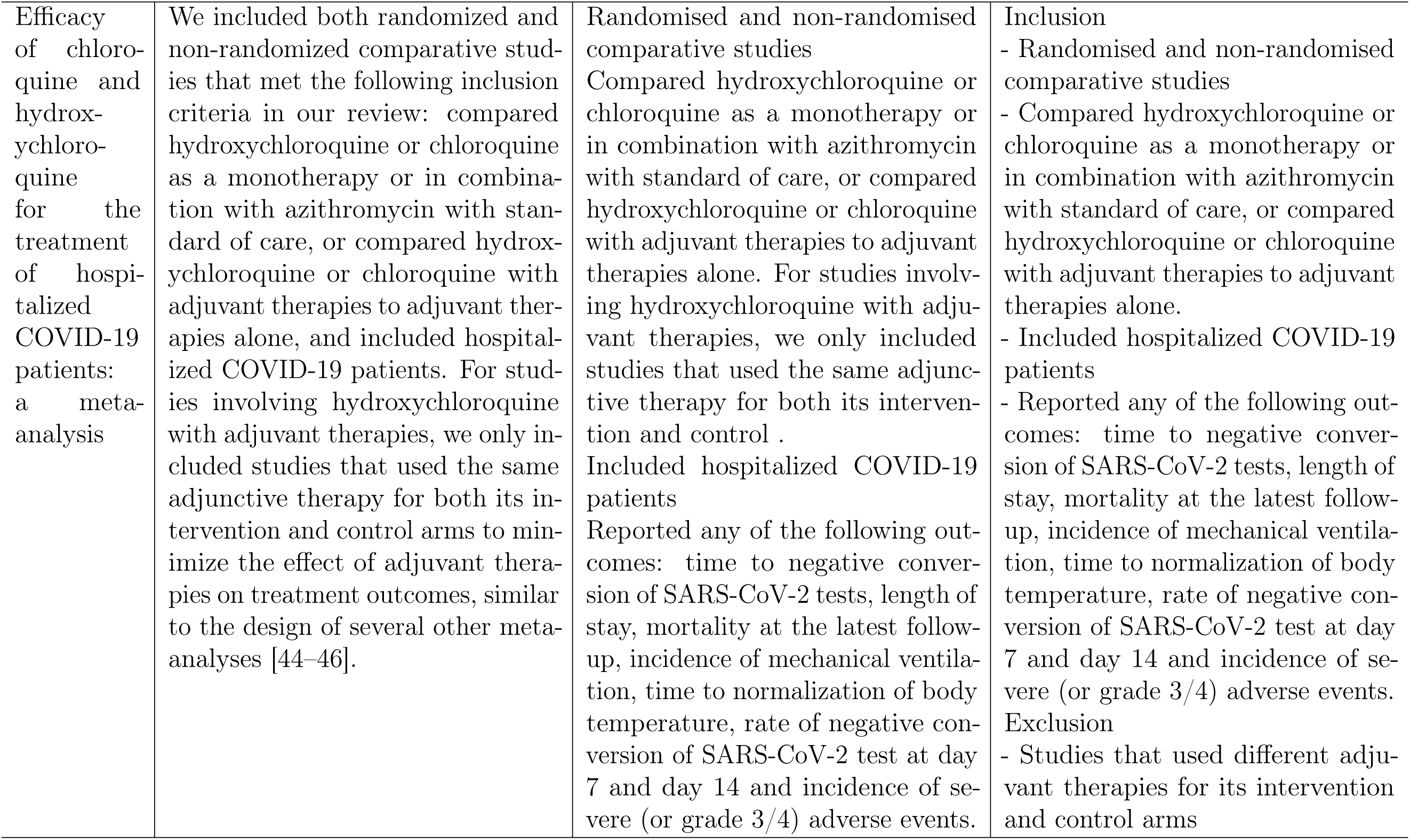
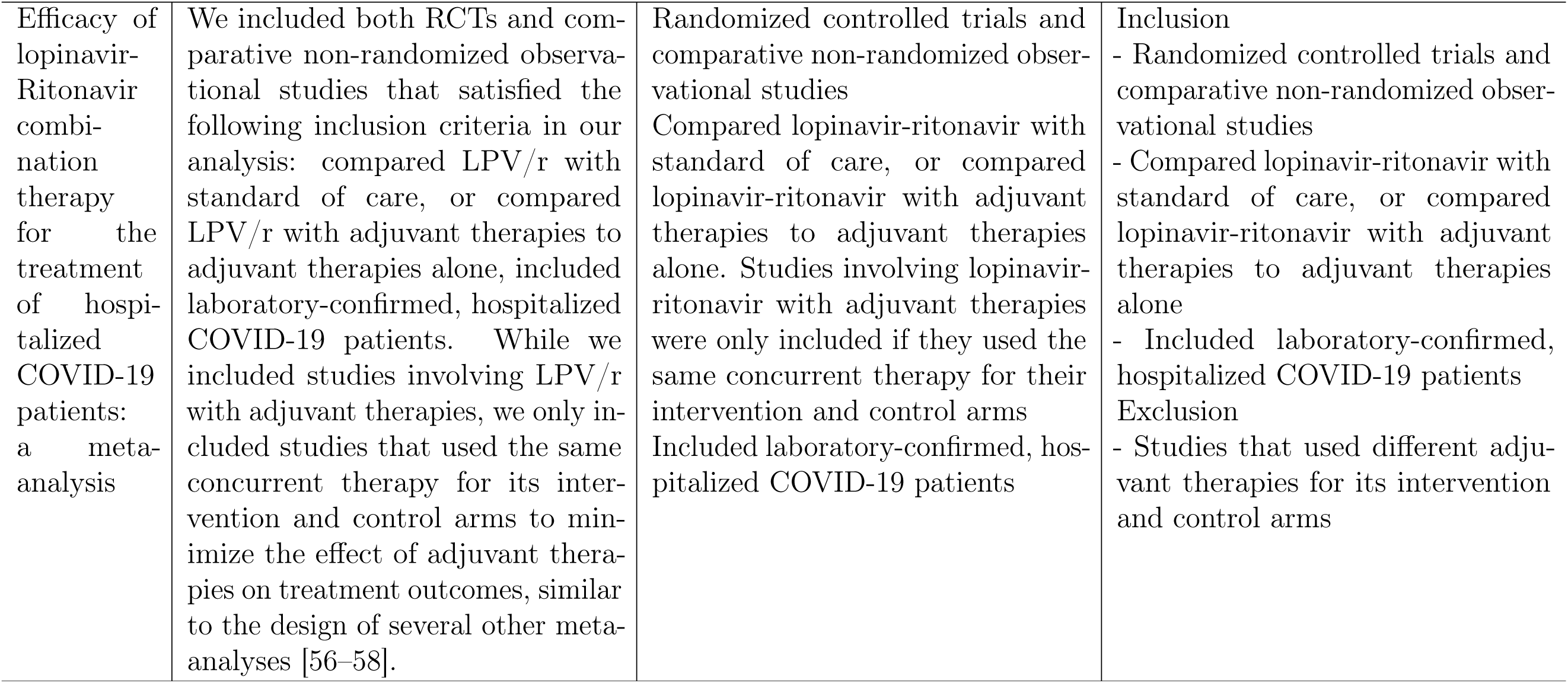
Criteria for COVID-19 topics.

| Topic | Free Text Criteria | Structured Criteria | Inclusion/Exclusion Criteria |
| --- | --- | --- | --- |
| Efficacy and safety of ivermectin for the treatment of COVID-19: a systematic review and meta-analysis | We included both randomized and non-randomized comparative studies that met the following criteria: (i) compared ivermectin to standard of care or a control group receiving placebo; (ii) included adult COVID-19 inpatients and/or outpatients; and (iii) reported any of our outcomes of interest. | Randomized controlled trials and non-randomized comparative cohort studies<br>Compared ivermectin against standard of care or control group/placebo<br>Included adult (age $\geq 18$) COVID-19 inpatients and/or outpatients. Studies that used ivermectin for prophylaxis of COVID-19 were excluded.<br>Reported any of the following outcomes: time to viral clearance, duration of hospitalization, mortality incidence, incidence of progression to mechanical ventilation, all-cause adverse events, incidence of investigator-defined serious adverse events<br>Peer-reviewed articles only | Inclusion<br>- Randomized controlled trials and non-randomized comparative cohort studies<br>- Compared ivermectin against standard of care or control group/placebo<br>- Included adult (age $\geq 18$) COVID-19 inpatients and/or outpatients<br>- Reported any of the following outcomes: time to viral clearance, duration of hospitalization, mortality incidence, incidence of progression to mechanical ventilation, all-cause adverse events, incidence of investigator-defined serious adverse events<br>Exclusion<br>- Studies that used ivermectin for prophylaxis of COVID-19<br>- Non-peer-reviewed articles |
| Clinical efficacy and safety of SARS-CoV-2-neutralizing monoclonal antibody in patients with COVID-19: A living systematic review and meta-analysis | Articles that met the following requirements were included in the study: 1) patients were adults with COVID-19; 2) interventions using neutralizing mAbs; 3) the comparator was a placebo or standard of care treatment; and 4) outcomes included primary or secondary outcomes (the primary outcomes included all-cause mortality at 28 days and hospitalization or hospital visit. Clinical recovery, ICU admission, mechanical ventilation, hospital discharge, and serious adverse events were classified as secondary outcomes.); 5) The study was a RCT. Only English and Korean studies were included in this meta-analysis. Two review authors independently and in duplicate evaluated publications for inclusion based on title and abstract and then reviewed relevant full-text articles. Disagreements during the review process were addressed by consensus, with the involvement of a third review author. | Randomised controlled trials<br>Compared interventions using neutralizing monoclonal antibodies (Etesevimab, bamlanivimab, casirivimab, imdevimab, casirivimab, imdevimab, bamlanivimab, etesevimab, sotrovimab, bebtelovimab, tixagevimab, cilgavimab) with a placebo or standard of care treatment<br>Included adults with COVID-19<br>Reported any of the following outcomes: all-cause mortality at 28 days and hospitalization or hospital visit. Clinical recovery, ICU admission, mechanical ventilation, hospital discharge, and serious adverse events<br>Articles in English or Korean | Inclusion<br>- Randomised controlled trials<br>- Compared interventions using neutralizing monoclonal antibodies with a placebo or standard of care treatment<br>- Included adults with COVID-19<br>- Reported any of the following outcomes: all-cause mortality at 28 days and hospitalization or hospital visit. Clinical recovery, ICU admission, mechanical ventilation, hospital discharge, and serious adverse events<br>- Articles in English or Korean |
| Efficacy and safety of corticosteroid regimens for the treatment of hospitalized COVID-19 patients: a meta-analysis | We included both randomized and non-randomized comparative studies that met the following inclusion criteria: compared any corticosteroid therapy with standard of care, or compared any corticosteroid therapy with an adjuvant therapy to adjuvant therapy alone, and included hospitalized COVID-19 patients. We only included studies using adjuvant therapies if the adjuvant therapy between the control and experimental arms are the same in order to minimize the effect of the adjuvant therapies on treatment outcomes. | Randomised and non-randomised comparative studies<br>Compared any corticosteroid therapy with standard of care, or compared any corticosteroid therapy with an adjuvant therapy to adjuvant therapy alone. We only included studies using adjuvant therapies if the adjuvant therapy between the control and experimental arms are the same in order to minimize the effect of the adjuvant therapies on treatment outcomes.<br>Included hospitalized COVID-19 patients<br>Reported any of the following outcomes: time to negative conversion of SARS-CoV-2 tests, overall length of hospitalization, length of ICU hospitalization, incidence of progression to ICU hospitalization, incidence of mortality, incidence of progression to mechanical ventilation, length of mechanical ventilation, incidence of all-cause adverse events, incidence of investigator-defined severe adverse events, incidence of hyperglycemia or corticosteroid-induced diabetes mellitus and incidence of nosocomial bacterial or fungal infections. | Inclusion<br>- Randomised and non-randomised comparative studies<br>- Compared any corticosteroid therapy with standard of care, or compared any corticosteroid therapy with an adjuvant therapy to adjuvant therapy alone.<br>- Included hospitalized COVID-19 patients<br>- Reported any of the following outcomes: time to negative conversion of SARS-CoV-2 tests, overall length of hospitalization, length of ICU hospitalization, incidence of progression to ICU hospitalization, incidence of mortality, incidence of progression to mechanical ventilation, length of mechanical ventilation, incidence of all-cause adverse events, incidence of investigator-defined severe adverse events, incidence of hyperglycemia or corticosteroid-induced diabetes mellitus and incidence of nosocomial bacterial or fungal infections.<br>Exclusion<br>- Studies that used different adjuvant therapies for its intervention and control arms |
| Efficacy and safety of selective serotonin reuptake inhibitors in COVID-19 management: a systematic review and meta-analysis | We included RCTs and prospective or retrospective observational studies that included inpatients and/or outpatients of any age with COVID-19 and compared the use of SSRIs to the use of the standard of care or placebo. Studies assessing patients with past or ongoing SSRI prescriptions at the time of diagnosis were specifically excluded. We did not impose any language restrictions. | Randomized controlled trials and non-randomized comparative cohort studies<br>Included COVID-19 inpatients and/or outpatients of any age<br>Compared the use of selective serotonin reuptake inhibitors (mainly fluvoxamine, fluoxetine, and citalopram/escitalopram) against standard of care or placebo<br>Studies assessing patients with past or ongoing prescriptions of selective serotonin reuptake inhibitors at the time of COVID-19 diagnosis were excluded.<br>Reported any of the following outcomes: all-cause mortality, hospitalization, composite outcome of emergency room visits or hospitalization, hypoxemia, requirement for supplemental oxygen, requirement for ventilator support, serious adverse events | Inclusion<br>- Randomized controlled trials and non-randomized comparative cohort studies<br>- Included COVID-19 inpatients and/or outpatients of any age<br>- Compared the use of selective serotonin reuptake inhibitors (mainly fluvoxamine, fluoxetine, and citalopram/escitalopram) against standard of care or placebo<br>- Reported any of the following outcomes: all-cause mortality, hospitalization, composite outcome of emergency room visits or hospitalization, hypoxemia, requirement for supplemental oxygen, requirement for ventilator support, serious adverse events<br>Exclusion<br>- Studies assessing patients with past or ongoing prescriptions of selective serotonin reuptake inhibitors at the time of COVID-19 diagnosis |
| Efficacy of chloroquine and hydroxychloroquine for the treatment of hospitalized COVID-19 patients: a meta-analysis | We included both randomized and non-randomized comparative studies that met the following inclusion criteria in our review: compared hydroxychloroquine or chloroquine as a monotherapy or in combination with azithromycin with standard of care, or compared hydroxychloroquine or chloroquine with adjuvant therapies to adjuvant therapies alone, and included hospitalized COVID-19 patients. For studies involving hydroxychloroquine with adjuvant therapies, we only included studies that used the same adjunctive therapy for both its intervention and control arms to minimize the effect of adjuvant therapies on treatment outcomes, similar to the design of several other meta-analyses [44–46]. | Randomised and non-randomised comparative studies<br>Compared hydroxychloroquine or chloroquine as a monotherapy or in combination with azithromycin with standard of care, or compared hydroxychloroquine or chloroquine with adjuvant therapies to adjuvant therapies alone. For studies involving hydroxychloroquine with adjuvant therapies, we only included studies that used the same adjunctive therapy for both its intervention and control .<br>Included hospitalized COVID-19 patients<br>Reported any of the following outcomes: time to negative conversion of SARS-CoV-2 tests, length of stay, mortality at the latest follow-up, incidence of mechanical ventilation, time to normalization of body temperature, rate of negative conversion of SARS-CoV-2 test at day 7 and day 14 and incidence of severe (or grade 3/4) adverse events. | Inclusion<br>- Randomised and non-randomised comparative studies<br>- Compared hydroxychloroquine or chloroquine as a monotherapy or in combination with azithromycin with standard of care, or compared hydroxychloroquine or chloroquine with adjuvant therapies to adjuvant therapies alone.<br>- Included hospitalized COVID-19 patients<br>- Reported any of the following outcomes: time to negative conversion of SARS-CoV-2 tests, length of stay, mortality at the latest follow-up, incidence of mechanical ventilation, time to normalization of body temperature, rate of negative conversion of SARS-CoV-2 test at day 7 and day 14 and incidence of severe (or grade 3/4) adverse events.<br>Exclusion<br>- Studies that used different adjuvant therapies for its intervention and control arms |
| Efficacy of lopinavir-Ritonavir combination therapy for the treatment of hospitalized COVID-19 patients: a meta-analysis | We included both RCTs and comparative non-randomized observational studies that satisfied the following inclusion criteria in our analysis: compared LPV/r with standard of care, or compared LPV/r with adjuvant therapies to adjuvant therapies alone, included laboratory-confirmed, hospitalized COVID-19 patients. While we included studies involving LPV/r with adjuvant therapies, we only included studies that used the same concurrent therapy for its intervention and control arms to minimize the effect of adjuvant therapies on treatment outcomes, similar to the design of several other meta-analyses [56–58]. | Randomized controlled trials and comparative non-randomized observational studies<br>Compared lopinavir-ritonavir with standard of care, or compared lopinavir-ritonavir with adjuvant therapies to adjuvant therapies alone. Studies involving lopinavir-ritonavir with adjuvant therapies were only included if they used the same concurrent therapy for their intervention and control arms<br>Included laboratory-confirmed, hospitalized COVID-19 patients | Inclusion<br>- Randomized controlled trials and comparative non-randomized observational studies<br>- Compared lopinavir-ritonavir with standard of care, or compared lopinavir-ritonavir with adjuvant therapies to adjuvant therapies alone<br>- Included laboratory-confirmed, hospitalized COVID-19 patients<br>Exclusion<br>- Studies that used different adjuvant therapies for its intervention and control arms |

## Footnotes

1 Llama: https://huggingface.co/meta-llama

2 https://github.com/automation-in-systematic-reviews/M-PreSS

